# FRONTLINE COMMUNITIES AND SARS-COV-2 - MULTI-POPULATION MODELING WITH AN ASSESSMENT OF DISPARITY BY RACE/ETHNICITY USING ENSEMBLE DATA ASSIMILATION

**DOI:** 10.1101/2021.02.27.21252589

**Authors:** Emmanuel Fleurantin, Christian Sampson, Daniel Paul Maes, Justin Bennet, Tayler Fernandez-Nunez, Sophia Marx, Geir Evensen

## Abstract

The COVID-19 pandemic has imposed many strenuous effects on the global economy, community, and medical infrastructure. Since the outbreak, researchers and policymakers have scrambled to develop ways to identify how COVID-19 will affect specific sub-populations so that good public health decisions can be made. To this end, we adapt the work of Evensen *et al* [1] which introduces a SEIR model that incorporates an age-stratified contact matrix, a time dependent effective reproduction number *R*, and uses ensemble data assimilation to estimate model parameters. The adaptation is an extension of Evensen’s modeling framework, in which we model sub-populations with varying risks of contracting SARS-CoV-2 (the virus that causes COVID-19) in a particular state, each with a characteristic age-stratified contact matrix. In this work, we will focus on 9 U.S. states as well as the District of Columbia. We estimate the effective reproductive number as a function of time for our different sub-populations and then divide them into two groups: frontline communities (FLCs) and the complement (NFLCs). Our model will account for mixing both within populations (intra-population mixing) and between populations (inter-population mixing). Our data is conditioned on the daily numbers of accumulated deaths for each sub-population. We aim to test and demonstrate methodologies that can be used to assess critical metrics of the pandemic’s evolution which are difficult to directly measure. The output may ultimately be of use to measure the success or failures of the pandemic response and provide experts and policymakers a tool to create better plans for a future outbreak or pandemic. We consider the results of this work to be a reanalysis of pandemic evolution across differently affected sub-populations which may also be used to improve modeling and forecasts.

## 1. Introduction

The COVID-19 pandemic has amplified social and economic inequities that impact the wellness of racial and ethnic minority groups. Such inequities increase the risk of infection and death for communities of color and other groups affected by systemic racism and economic inequality. Our goal is to develop a multi-population framework that can be used to study how the virus spreads in different populations, detect inequities, and provide estimates of crucial metrics that can be used to understand any disparities. To this end, we consider COVID- 19 data and population statistics from the states of Alaska, California, Connecticut, Delaware, Hawaii, Maryland, Michigan, Utah, and Washington, as well as the District of Columbia, on account of their more complete reporting of deaths by race/ethnicity. In particular, for exploratory purposes, we choose to look at different populations that can all be categorized as frontline communities (FLCs) to comprise our multiple populations [2, 3, 4, 5, 6, 7]. We aim to use iterative ensemble smoothers [8, 9], often used in geosciences [10] or in petroleum reservoir modeling [11], to estimate parameters for a multi-population SEIR model with age-classes and compartments representing hospitalized, sick, and dead individuals.

We have extended Evensen *et al*.’s model [1] to study the evolution of the COVID- 19 pandemic and have subsequently used it to perform a set of simulations for different populations in the regions previously mentioned. The system revises the model state and calibrates its parameters to fit a time series of indirect and noisy observations of deaths, hospitalizations, and infected individuals [1]. Here for reasons explained below we focus on only deaths for assimilation. Different socio-economic conditions among these groups have led the epidemic to evolve differently among such populations and thus have different impacts on them. Moreover, the counter- measures implemented by certain states have affected the way we study how the virus spreads within different populations as well. Matters are further complicated by the scarcity of COVID-19 data for different ethnic and racial groups, especially minority groups, within a particular state.

We believe that data assimilation is very important here in that the model will always track the data to a degree of certainty that is proportional to its assumed accuracy, enabling a straightforward treatment of incomplete and noisy data [12]. The use of ensemble data assimilation helps us understand the impact of the implemented measures on the magnitude of the effective reproductive number, *R*_*t*_(*n*), as a function of time, *t*, on a population, *n*. Moreover, with knowledge of daily infections (tested) and deaths due to COVID-19 for different racial/ethnic groups, we make inferences on how the disease has affected different sub-populations under distinct social distancing scenarios. Additionally, using iterative ensemble smoothers is beneficial in this case since state and parameter estimations are done by constraining the parameters on all the data in a given time window [9].

Some notable works have used data assimilation for epidemiology, in the context of both variational [13] and Kalman filter-like methods [14]. Most of the studies use sequential filtering approaches, e.g., the iterative filter and the ensemble Kalman filter (EnKF) or the Ensemble Adjustment Kalman Filter (EAKF). Recent studies on SARS-CoV-2 still mostly use a filtering approach. A filter will update bias in the solution. However, it does not adjust the parameters that can lead to that bias. Instead of filtering, we make use of the ensemble data-assimilation method (ESMDA) which acts as a smoother, updating the bias in the parameters rather than just in the solution alone. This is critical in the estimation of *R*_*t*_(*n*) since adjustments to the value of it at a given time are adjusted to predict death data about two weeks later. In this way, we are updating the parameters in the past to remove the bias in the solution before it happens.

The analysis resulting from this approach can be used to help public health officials understand certain aspects of the COVID-19 pandemic within specific sub-populations, make informed predictions regarding its course, and create intervention plans that better support disadvantaged groups. In particular, given the current dilemma surrounding the reopening of businesses, our model can help state officials understand the effects of population mixing between different age classes and help local officials analyze how different intervention strategies, such as partial or complete school closings, or work-from-home arrangements, can alter or even potentially help curtail the virus’ spread through frontline communities. The model-system code for our multi-population model is available from Github, https://github.com/geirev/EnKF_seir, and all plots were generated using MATLAB.

The outline of the paper is as follows: Section 1.1 provides our working definition of a frontline community, Section 2 describes the SEIR model used, and in Section 3 we give a brief introduction to the use of ensemble methods for model calibration. In Section 4 we highlight the model behavior between a frontline community (FLC) and non-frontline community (NFLC). We describe how we choose their representative age-stratified contact matrices and provide the general setup for our ESMDA runs, presenting individual results that we discuss for the different states we consider. In Section 5 we conclude by making an overall assessment of the results obtained across the modeled populations within the 9 states previously mentioned and the District of Columbia, as well as presenting some next steps.

### 1.1 Working Definition of a Frontline Community (FLC)

In this work, we consider “frontline communities” (FLCs) to be subgroups of the overall population that encompass a multitude of different socio-economic roles and factors. In general, such factors reflect apparent levels of inequality and inequity available to such subgroups of the population. For example, certain racial/ethnic groups for a given region could account for a disproportionate representation of jobs that are considered more high-risk for contracting COVID-19 (e.g. retail/service/health care workers). Another factor of interest could be those subgroups of the population that have less total and/or inter-generational wealth than others or who are ineligible for receiving federal and state assistance. This might affect how members of such groups could absorb or recover from the adverse economic effects of COVID-19 lockdowns and quarantines, in terms of both economic resilience and ability to maintain social isolation in high-density living situations. It may also be that an FLC is one that tends to be compelled into becoming a majority population in an underserved area in terms of hospitals and general medical infrastructure.

Overall, FLCs share a commonality in that they are subgroups that are disproportionately negatively impacted by the spread of COVID-19. For this work, we will adopt the same criteria used in [2] where different criteria are used to identify a group for likely disparity. A group is flagged for likely disparity when case counts or deaths meet the following criteria: (1) is at least 33% higher than the Census Percentage of Population, (2) remains elevated whether including or excluding cases/deaths with unknown race/ethnicity, (3) is based on at least 30 actual cases or deaths. It is important to note that these criteria can still fail to detect disparities. For example, one group may be flagged as an FLC in terms of confirmed cases while another may not and goes without notice due to lack of testing or case reporting for that subgroup. Using ESMDA techniques we can estimate the actual infections in a group through the assimilation of the death data and detect some of these kinds of possible disparities. Indeed, as is discussed in Section 4.2.3, for the state of Connecticut we see that while the Latinx community is flagged for the disparity in terms of confirmed cases, we find that the Black community may in fact make up more of the total infections despite being 6% less of the total population. The Black community, however, was not flagged as having a percentage of confirmed cases compared to the percentage of the total population.

In total there are 9 possible racial/ethnic groups reported in the data, White, Black, Latinx, American Indian and Alaskan Native (AIAN), Asian, Native Hawaiian and Pacific Islander (NHPI), Multiple, Other and Unknown. Each locality that we study may report some of these groups differently or not at all, and in some cases they may even be double counted. The Unknown group is more a measure of deaths not assigned to any group and is not considered other than to gauge uncertainty as it is not a racial/ethnic group.

## 2. Model

### 2.1 Evensen *et al*. Original (One Population) Model

In the original Evensen *et al*. model, there are 3*n*_*a*_ + 9 compartments–and a corresponding set of differential equations–which describe the spread of the disease for a population split into *n*_*a*_ age groups. There are *n*_*a*_ compartments/equations for susceptible (*S*_*i*_), exposed (*E*_*i*_), and infectious (*I*_*i*_) individuals in each age group, with *i* = 1, *…, n*_*a*_. The remaining compartments and equations are for the following groups: quarantined individuals with mild (*Q*_*m*_), severe (*Q*_*s*_), or fatal (*Q*_*f*_) symptoms; hospitalized individuals with severe (*H*_*s*_) or fatal (*H*_*f*_) symptoms; individuals with fatal symptoms who are put into a care home (*C*_*f*_); recovered individuals who had mild (*R*_*m*_) or severe (*R*_*s*_) symptoms; and dead individuals (*D*).

An infectious individual from the age group *j* (*I*_*j*_) can infect a susceptible individual in any age group *i* (*S*_*i*_). They will do so at a rate *R*_*ij*_(*t*), which highlights the different contact/infection rates between each age group. These values compose the matrix 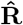. We also have a scalar function *R*(*t*) (note that this is equivalent to the function *R*_*t*_(*n*) mentioned previously with *n* = 1 and with simplified notation) which is the effective reproductive number at time *t*. Multiplying these two quantities together gives 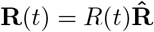. We can ensure that *R*(*t*) solely determines the effective reproductive at time *t* by weighting 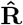 such that 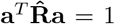, where **a** is a vector, with elements *a*_*i*_ that are the proportion of the population comprising age group *i*. The system runs through three different intervention periods: (1) “pre-lockdown” (before any mitigation measures), (2) “lockdown” (when people were asked or required to stay home and avoid gathering), and (3) “post-lockdown” (when a state began to reopen businesses and lessen restrictions on gatherings). For each of these time periods, we can have different 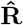 matrices, 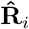 to reflect different contact rates between different age groups in each time period.

We have the following parameters for this model: the proportion of the population that has mild symptoms and is from age group *i* 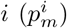, the proportion of the population that has severe symptoms, and is from age group *i* 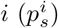, the proportion of the population that has fatal symptoms and is from age group *i* 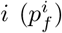, the proportion of infected individuals with fatal symptoms who are hospitalized (*p*_*h*_), the average length of the incubation period (*τ*_inc_), the average length of the infectious period (*τ*_inf_), the average length of hospitalization (*τ*_hosp_), the average recovery time for individuals with mild symptoms (*τ*_recm_), the average recovery time for individuals with severe symptoms (*τ*_recs_), and the average time to death for individuals with fatal symptoms (*τ*_death_).

### 2.2 Multi-Population Model Extension

#### 2.2.1 New Model Assumptions & Clarifications

Now, we employ a modified version of the original Evensen *et al*. model, extending it from a single population of interest to *n*_c_ populations and *n*_a_ age classes within each population. Here, different populations could represent different countries, states, localities, or specific population groups within a locality. For this work we consider each population to be the different racial/ethnic communities in a state. In the equations below we use the following notation: *n, m* are indices running over all *n*_c_ communities and *i, j* are indices running over all *n*_a_ age groups within in each community. The total populations of any two communities, *n* and *m*, are given by *N*_*n*_ and *N*_*m*_.

We model the interaction between groups using the elements of 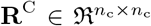 (the diagonal must always be 1) and 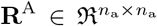. The effective reproductive number per community is a scalar function of time, *R*_*t*_(*n*), and is a parameter that we estimate. We model the relative differences in infectiousness between age groups using the coefficients in 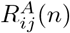, which can differ between communities. The model default is “null”, such that all elements are set to 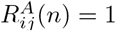, which assumes equal transmission rates among all age groups. In the equations below, we use the 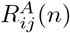 for age group *i* as it interacts with age group *j*. The only sound alternative to this choice would be to set 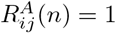 when *m ≠ n*, given the number of coefficients we would otherwise need to specify. An example of such a matrix 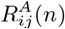 can be seen in Figure 1.

**Figure 1.**
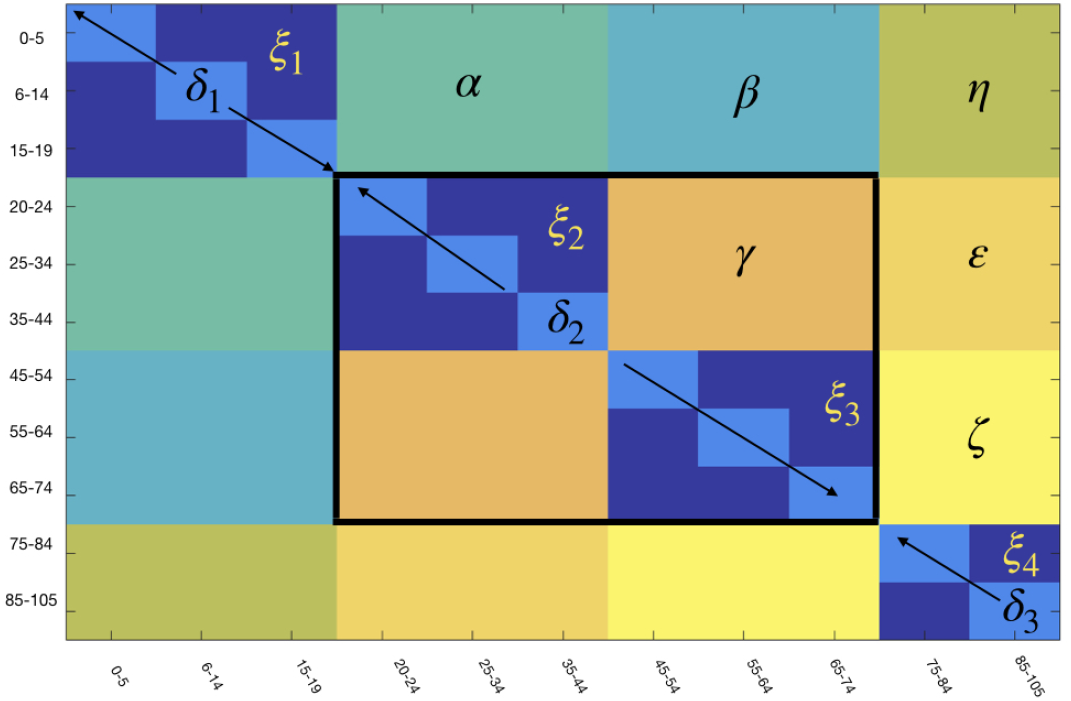
The general form of the 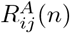 contact matrix elements for contact rates between age groups *i* and *j* in a given sub-population. The contact matrix is then subdivided into different blocks where parameters *α, β, η, γ, ε, ζ, δ*_1_, *δ*_2_, *δ*_3_, *ξ*_1_, *ξ*_2_, *ξ*_3_ and *ξ*_4_ control the contact rates between different age groups which generate similar patterns for spreading the disease. In particular, we define *γ, δ*_2_, *ξ*_2_ and *ξ*_4_ to be the parameters for the contact rates of the working class age groups.

The fractions of mildy, fatally, and severely ill (*p*_m_, *p*_f_, and *p*_s_, respectively) can differ between communities. We have used the same hospitalization fraction of fatally ill, *p*_h_, for all communities.

In the case with only one community *n* = *m* = 1, we have *N*_*m*_*/N*_*m*_ = 1, as well as **R**^C^(*n, m*) = 1. Thus, the equations reduce to the standard SEIR model as in Section 2.1. The use of a multi-compartment model only changes the nonlinear interaction term present in the **S**_*i*_(*n*) and **E**_*i*_(*n*) equations. Outside of this interaction term, each community evolves independently of every other.

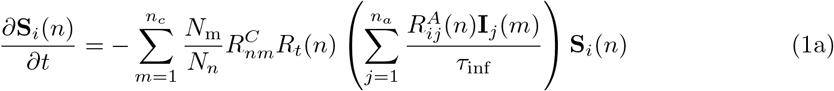

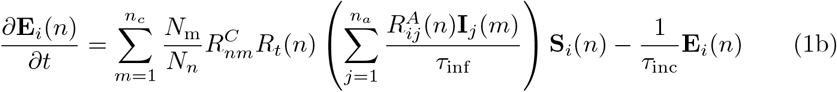

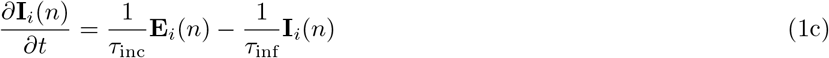

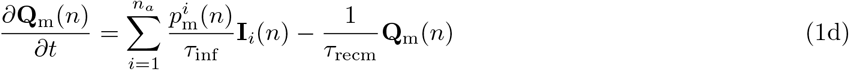

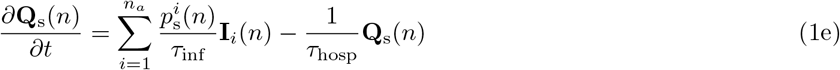

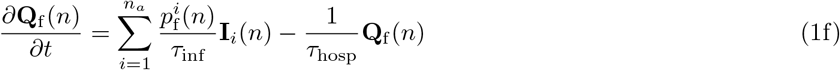

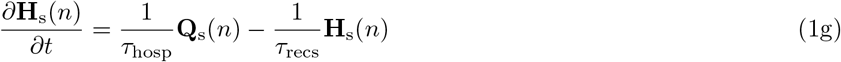

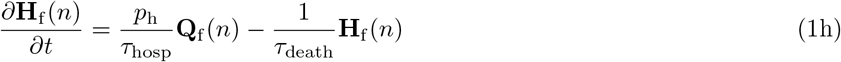

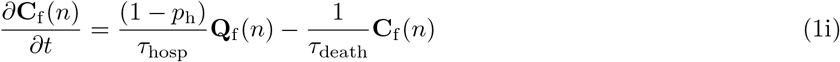

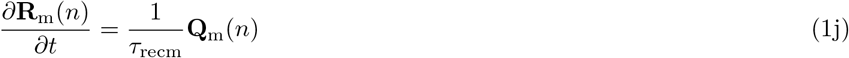

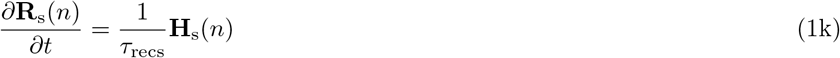

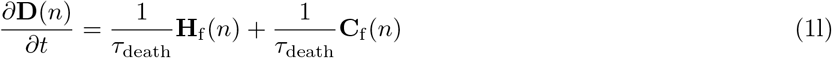

## 3. Methodology

A common issue with solving high-dimensional models is the difficulty in accurately estimating parameters. However, optimal estimates can be found using data assimilation methods. In particular, we highlight the use of an ensemble Kalman Filter (EnKF method) for sequential data assimilation. In recent history, ensemble data assimilation has been widely implemented in weather predicting [15], tumor growth and spread [16], and petroleum reservoir history matching [11]. The existing theory and application in these fields have seen the implementation of ensemble data assimilation in other fields, such as epidemiology. In particular, we take inspiration from Evensen *et al*.’s use of an ensemble smoother with multiple data assimilation (ESMDA) in their extended SEIR model of COVID-19 upon which we build [1].

### 3.1 The inverse problem

Ensemble smoother techniques can be derived by assuming a perfect forward model.

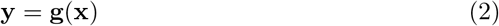

In general, **x** is the realization of model parameters, and **y** consists of the uniquely predicted measurements. For the case of COVID-19, **x** consists of the initial conditions, parameters, and time-reliant effective reproductive numbers. We relate the predictions **y**, to the parameters **x** through the model operator **g**(**x**), where **g** is the model in Equation 1.

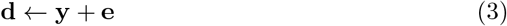

In our model, **y** then consists of the predicted measurements of deaths, cases and hospitalizations given some model error, **e**. Here, **d** is the observed data. To solve the inverse problem, it is efficient to frame it into an equation using Bayes’ theorem:

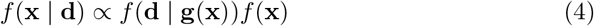

Equation (4) represents the so-called smoothing problem, which can be approximated using ensemble methods. We refer the reader to Evensen’s analysis of solving inverse problems for a full derivation and well-constructed analysis in [17].

### 3.2 ESMDA

The assimilation method used is an iterative ensemble smoothing method called an ensemble smoother with multiple data assimilation. The method, which is similar to an ensemble smoother, solves the parameter-estimation problem and is formally derived from the Bayesian formulation using a tempering procedure [18]. What separates this method from other similar methods is that it approximates the posterior recursively, gradually introducing information to alleviate the impact of nonlinear approximation. After updating through to the last time step, it begins the assimilation process over again - resampling the vector of perturbed observations, which reduces sampling error [19].

Even though we focus on ensemble smoothing conceptually, the procedure can also be applied to ensemble Kalman filtering. Multiple data assimilation gives only small improvements when applied to an ensemble Kalman filter, which makes ESMDA a much more effective option in terms of computational cost [17]. The simplicity and effectiveness of ESMDA are what make it an optimal assimilation method for this particular application.

For simplicity, we lay out the ensemble methods without the mathematical details, as laid out in [1].

- First, sample a large ensemble of realizations of the prior uncertain parameters (age groups, the functions *R*_*t*_(*n*), and the initial infected and exposed), given their prescribed first-guess values and standard deviations.
- Integrate the ensemble of model realizations forward in time to produce a prior ensemble prediction, which also characterizes the uncertainty.
- Compute the posterior ensemble of parameters by using the misfit between prediction and observations, and the correlations between the input parameters and the predicted measurements.
- Finally, compute the posterior ensemble prediction by a forward ensemble integration. The posterior ensemble is then the “optimal” model prediction with the ensemble spread representing the uncertainty.

## 4. Case Studies

In this section, we highlight how the model behaves when using different age-stratified matrices for two populations, one FLC and one NFLC population. During the different intervention periods, we make the assumption that FLCs will have increased contact rates to varying degrees across all age groups. The matrices are described below. We then describe our assumptions and various approaches to the ESMDA problem and interpret the results obtained for each state in our study. First we make some remarks on the model parameters and data chosen for assimilation.

For the model parameters explained in Section 2 we use the same initial guesses as in [1] and show them in Tables 4 and 5 in Appendix A. The initial values for theses parameters are based off values obtained available data and some initial model tuning experiments. However, the DA will fine tune these parameters if necessary. We use the same initial parameters for all communities including the *p*-numbers and case fatality rates. While there is some evidence that different racial/ethnic communities may have more members with underlying conditions, this is also true of members of the same community who live in different regions. We do not find enough available data to make confident guesses in any differences in these parameters amongst the different communities or localities. We also believe the differences to be relatively small and that any discrepancies can be accounted for by a small increase or decrease in *R*_*t*_(*n*) as estimated by the ESMDA scheme. In general we find that for our best obtained results these initial parameters are good estimates and that *R*_*t*_(*n*) is the primary differentiator between communities. It should be noted though that some communities such as the AIAN community in Alaska may not have as much access to needed medical care in Arctic rural areas. This could mean a higher CFR and lower *R*_*n*_(*t*) is warranted, however the DA will still detect discrepancies through *R*_*n*_(*t*).

**Table 1.** Matrix scaling parameters for FLC and NFLC workers in lockdown and post-lockdown time periods.

| Scalings | Matrix Parameters |  |  |  |  |  |  |  |  |  |  |  |  |
| --- | --- | --- | --- | --- | --- | --- | --- | --- | --- | --- | --- | --- | --- |
| | $\alpha$ | $\beta$ | $\gamma$ | $\eta$ | $\epsilon$ | $\zeta$ | $\delta_1$ | $\delta_2$ | $\delta_3$ | $\xi_1$ | $\xi_2$ | $\xi_3$ | $\xi_4$ |
| FLC lock | 0.5 | 0.7 | 0.55 | 0.25 | 0.25 | 0.35 | 0.3 | 0.7 | 0.7 | 0.4 | 0.65 | 0.55 | 0.6 |
| FLC post-lock | 0.7 | 0.8 | 0.7 | 0.3 | 0.3 | 0.4 | 0.6 | 0.85 | 0.7 | 0.85 | 0.7 | 0.65 | 0.65 |
| NFLC lock | 0.5 | 0.7 | 0.5 | 0.2 | 0.2 | 0.3 | 0.3 | 0.6 | 0.7 | 0.4 | 0.6 | 0.5 | 0.6 |
| NFLC post-lock | 0.7 | 0.8 | 0.7 | 0.25 | 0.25 | 0.35 | 0.6 | 0.7 | 0.75 | 0.7 | 0.7 | 0.65 | 0.65 |

**Table 2.** Date breakdown by intervention periods for all states.

| Interventions | Information on Intervention Periods by State |  |  |  |  |  |  |  |  |  |
| --- | --- | --- | --- | --- | --- | --- | --- | --- | --- | --- |
|  | AK | CA | CT | DC | DE | HI | MD | MI | UT | WA |
| Start date | 3/8/20 | 2/25/20 | 2/27/20 | 2/27/20 | 3/1/20 | 2/28/20 | 2/25/20 | 2/21/20 | 2/29/20 | 1/9/20 |
| 1st Phase | 3/19/20 | 3/19/20 | 3/19/20 | 3/17/20 | 3/17/20 | 3/17/20 | 3/17/20 | 3/17/20 | 3/17/20 | 3/17/20 |
| 2nd Phase | 5/27/20 | 5/27/20 | 5/27/20 | 5/29/20 | 5/31/20 | 5/15/20 | 5/15/20 | 5/19/20 | 5/5/20 | 5/28/20 |

**Table 3.** Demographic breakdown by race/ethnicity (where data is available) for all states. Groups that meet the criteria to be an FLC are in bold. AIAN = American Indian and Alaska Native, NHPI = Native Hawaiian and Pacific Islander.

| Race/Ethnicity | Percentage of Population per State |  |  |  |  |  |  |  |  |  |
| --- | --- | --- | --- | --- | --- | --- | --- | --- | --- | --- |
|  | AK | CA | CT | DC | DE | HI | MD | MI | UT | WA |
| AIAN | **0.15** | 0.076 | X | X | X | X | X | 0.005 | 0.023 | **0.01** |
| Asian | 0.06 | 0.14 | 0.04 | 0.0435 | 0.045 | **0.38** | 0.06 | 0.035 | 0.038 | 0.08 |
| Black | 0.03 | 0.06 | 0.1 | **0.4453** | 0.22 | 0.02 | 0.29 | **0.14** | 0.021 | 0.04 |
| Latinx | X | **0.39** | **0.16** | **0.113** | **0.09** | X | **0.1** | X | 0.142 | **0.13** |
| Multi | X | X | **0.02** | X | 0.02 | X | X | X | X | 0.05 |
| NHPI | **0.01** | 0.039 | X | X | X | **0.1** | X | X | 0.016 | **0.008** |
| Other | 0.08 | X | 0.01 | 0.01 | X | 0.24 | X | 0.03 | 0.01 | 0.005 |
| White | 0.65 | 0.37 | 0.67 | 0.4196 | 0.62 | 0.25 | 0.51 | 0.78 | 0.78 | 0.69 |

**Table 4.** The table gives a set of first-guess model parameters. As we could not find scientific estimates of these parameters, we set their values based on available information from the internet and initial model-tuning experiments. We leave it to the data assimilation system to fine-tune the parameter values.

| Parameter | First guess | Description |
| --- | --- | --- |
| $\tau_{\text{inc}}$ | 5.5 | Incubation period |
| $\tau_{\text{inf}}$ | 3.8 | Infection time |
| $\tau_{\text{recm}}$ | 14.0 | Recovery time mild cases |
| $\tau_{\text{recs}}$ | 5.0 | Recovery time severe cases |
| $\tau_{\text{hosp}}$ | 6.0 | Time until hospitalization |
| $\tau_{\text{death}}$ | 16.0 | Time until death |
| $p_f$ | 0.009 | Case fatality rate |
| $p_s$ | 0.039 | Hospitalization rate (severe cases) |
| $p_h$ | 0.4 | Fraction of fatally ill going to hospital |

**Table 5.** The *p*-numbers indicate the fraction of sick people in an age group ending up with mild symptoms, severe symptoms (hospitalized), and fatal infection.

| Age group | 1 | 2 | 3 | 4 | 5 | 6 | 7 | 8 | 9 | 10 | 11 |
| --- | --- | --- | --- | --- | --- | --- | --- | --- | --- | --- | --- |
| Age range | 0–5 | 6–12 | 13–19 | 20–29 | 30–39 | 40–49 | 50–59 | 60–69 | 70–79 | 80–89 | 90–105 |
| Population | 351159 | 451246 | 446344 | 711752 | 730547 | 723663 | 703830 | 582495 | 435834 | 185480 | 45230 |
| p-mild | 1.0000 | 1.0000 | 0.9998 | 0.9913 | 0.9759 | 0.9686 | 0.9369 | 0.9008 | 0.8465 | 0.8183 | 0.8183 |
| p-severe | 0.0000 | 0.0000 | 0.0002 | 0.0078 | 0.0232 | 0.0295 | 0.0570 | 0.0823 | 0.1160 | 0.1160 | 0.1160 |
| p-fatal | 0.0000 | 0.0000 | 0.0000 | 0.0009 | 0.0009 | 0.0019 | 0.0061 | 0.0169 | 0.0375 | 0.0656 | 0.0656 |

For this work, we choose only to assimilate death data for each of the racial/ethnic communities. While the model and ESMDA scheme can assimilate case counts and hospitalizations, we find this data to currently be of too poor quality for assimilation. Case counts can be very misleading when the actual percentage of cases it captures is unknown and changing through time while hospitals may not report racial/ethnic data at all or quickly enough. However as deaths are recorded by each local government and each reports a deceased individual’s race, there is more consistency in the reporting. At the time of writing, the race/ethnic data set is somewhat incomplete for most states and is changing as things unfold. We choose states for which there is at least 93% reporting up to January 3, 2021 for our analysis. We also increase the uncertainty in the death data to account for unknown cases while keeping it low enough for the ESMDA to actually make informative updates. We believe more analysis should be done when the data is more complete and more data, such as hospitalizations, can be made readily available. However, we believe there is still much that can be learned in this analysis. The data that we use is compiled by The Covid Tracking Project who themselves compile the data from local government authorities [2].

### 4.1 Non-DA runs

We begin our simulations by working with two “toy” populations to see how the model performs with no DA. To that effect, we consider two populations with the following initial conditions: population 1 has an initial exposed of 500, initial infected of 350, and a case fatality ratio (CFR) of 0.009; population 2 has an initial exposed of 1000, initial infected of 700, and a CFR of 0.01. We use the following parameters for Equation 1 in our simulation, *τ*_inf_ = 3.8, *τ*_inc_ = 5.5, *τ*_recm_ = 14, *τ*_recs_ = 5, *τ*_hosp_ = 6, and *τ*_dead_ = 16. We use a continuous function for *R*_*t*_(*n*) with data from [20]. For this run, we use the time varying estimates of the effective reproductive number for the state of Utah obtained from [20] (see bottom left plot of Figures 3, 4). The interactions between age groups are taken into account in the 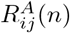 matrix entries (see Table 1 and Figure 2). Our decisions on the chosen values are based on assumptions that population 2 will be an FLC while population 1 will be an NFLC. We chose two scenarios with different proportions of the FLC to NFLC present in the total population.

**Figure 2.**
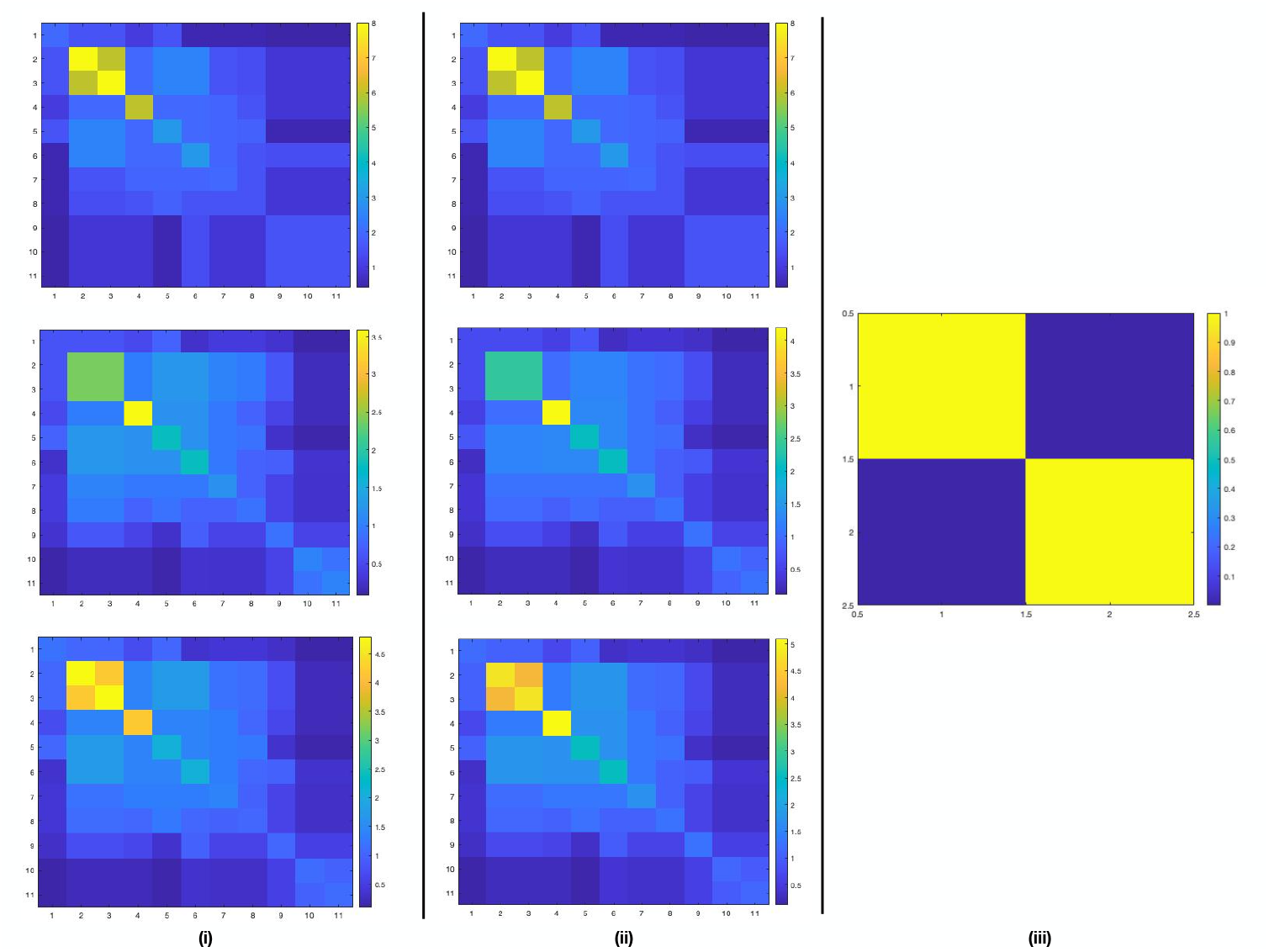
**R**^A^ for the NFLC (*i*) and FLC (*ii*) in our runs without DA for each intervention period along with the **R**^C^ matrix (*iii*) used in these simulations. We also used the **R**^A^’s in (*i*) and (*ii*) for the DA runs.

**Figure 3.**
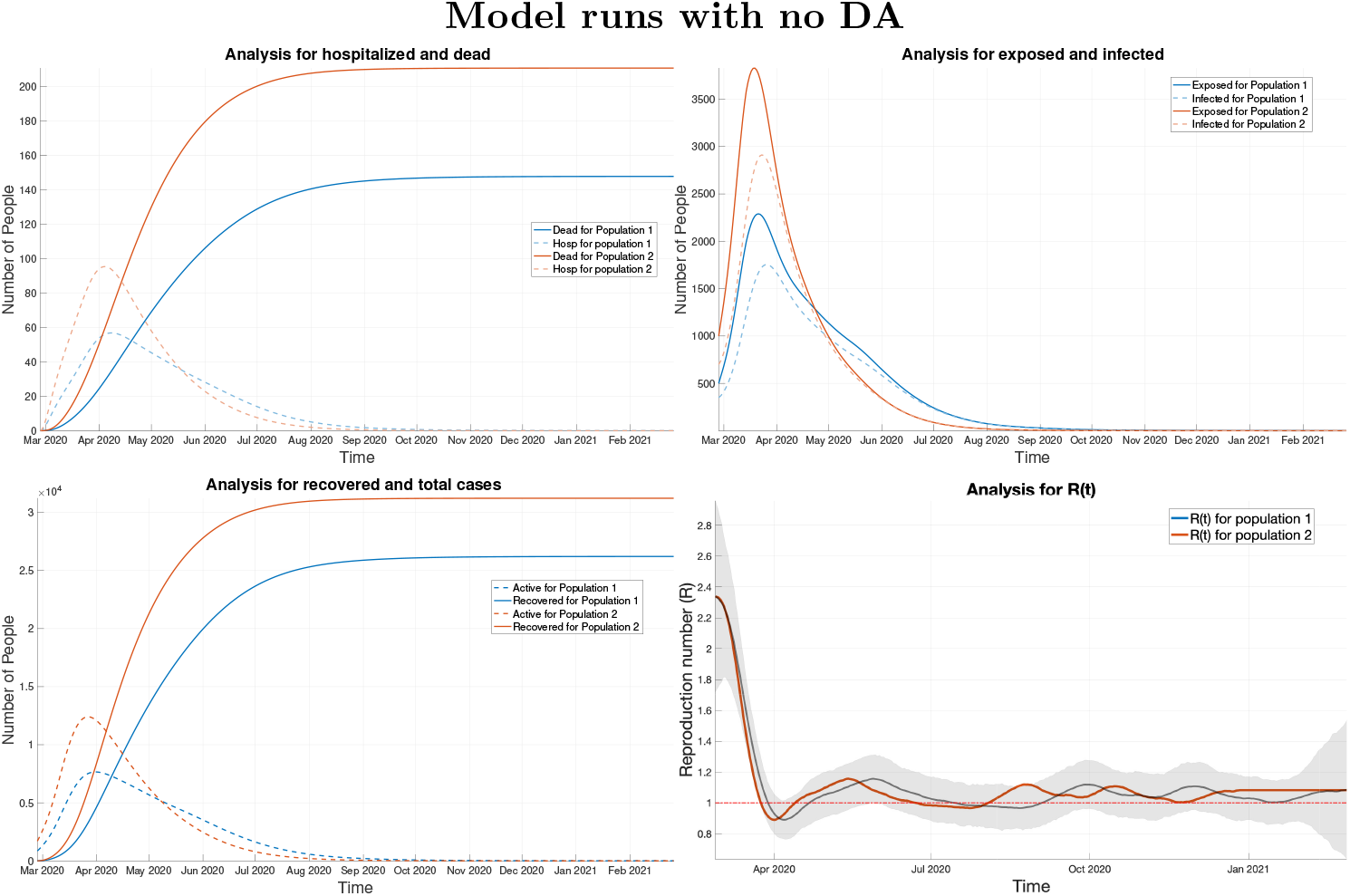
Results for non-DA runs with FLC and NFLC at about the same population.

**Figure 4.**
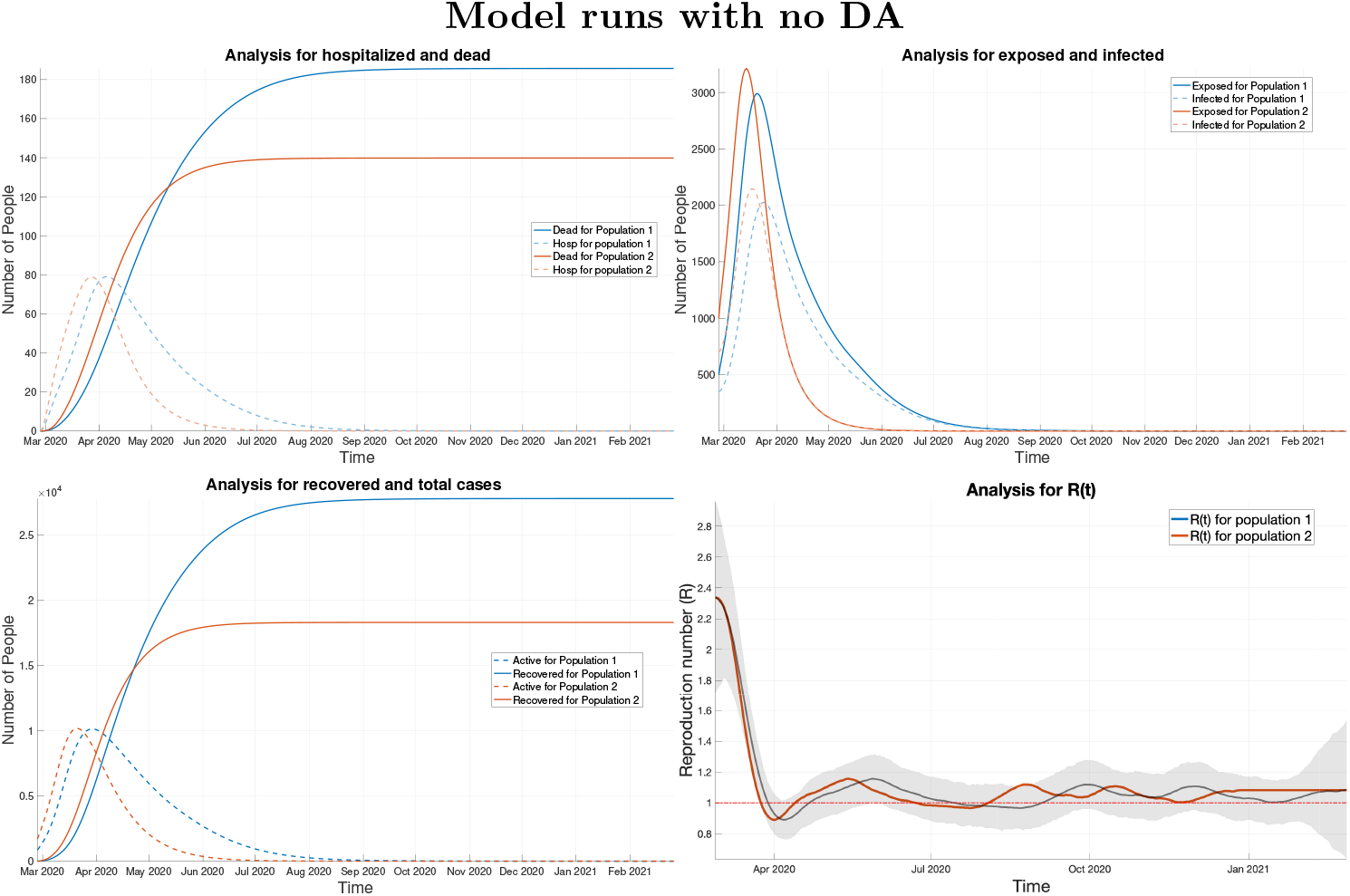
Results for non-DA runs with NFLC at 70% of the total population.

For the non DA runs, each matrix in (*i*), (*ii*) of Figure 2, is used to indicate the different “intervention periods” in the model: top =“pre-lockdown”, middle =“lockdown”, and bottom =“post-lockdown”); (*i*) represents contact matrices for NFLC workers and (*ii*) represents contact matrices for FLCs. For both (*i*) and (*ii*), the top matrices are the same. We used a contact matrix from a statistical survey in Europe [1] to describe the transmission between different age groups as a basis for such matrices. In Table 1 we note the parameter values that were used to scale the top matrix for each intervention period for both FLCs and NFLCs. During the lockdown intervention period, contact rates are decreased across all groups but more so for school-age children and older adults, in line with school closures and assumed caution amongst the most vulnerable populations. After a “re-opening” (i.e., entering the “post-lockdown” period), rates are increased across all age groups but more so for working-age adults and children, while assuming caution remains amongst the older population. For FLCs, contact rates between working adults and older adults are higher than for an NFLC. This builds in the assumption that FLC jobs tend to be in places where exposure is more likely (grocery stores, construction, etc) and that there are more inter-generational households connecting working-age adults and children with older adults in such communities. Identifying the scaling parameters is a difficult task and more data would be needed to estimate them. We take an ad-hoc approach to estimation but note that the normalization of 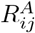 means that the primary control of the rate of spread is in the scaling factor *R*_*t*_(*n*) for the DA runs in Section 4.2. In (*iii*) we see the inter-population contract matrix between our two populations where the diagonal elements, 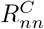, are 1 and the off-diagonal elements are very small (*<<* 1). The results show that the model takes into effect the total amount in each population as well as the contact matrices.

In Figure 3, both groups have about the same population. Since we are not assimilating data for these runs, we have only included the averages in the plots (which in this case is the same as if we were simply integrating a basic SEIR model). In all plots, the blue curves represent the NFLC while the red is the FLC. We can clearly note that the total amount of projected deaths and cases for the FLC tend to be higher since the intra-population contact rates for each intervention period are also higher for the frontline groups. In the bottom-left figure, we use data on the state of Utah as the effective reproductive number *R*_*t*_(*n*) for this simulation (black curve) retrieved from [20]; the shaded gray area around the curve represents the cone of uncertainty provided by the site. The blue and green curve represents *R*_*t*_(*n*) (they are the same in this case since we have no DA). In Figure 4, colors have the same meaning as in Figure 3. The NFLC represents a significant amount of the population (about 70%) in this scenario. The total amount of projected deaths and cases for the FLC is higher until June then projected deaths and cases for the NFLC surpasses it, but not by much.

### 4.2 DA Runs

Here we present the results of the ESMDA analysis performed on several states for which racial data reporting was at least above 93% [2]. We choose states which show very apparent disparities between groups (e.g. according to an analysis of disproportionate effects of COVID-19 on racial/ethnic groups in [2]) as well as some for which disparities are less apparent in the aggregate data. Initial conditions on the number of exposed and infected for each racial/ethnic community and state are estimated using data from The Institute for Health Metrics and Evaluation (IHME) [21] which provides an estimate of actual cases for each day of the pandemic. We take this estimate from the start date of our simulations and scale it by the proportion of the population each community represents. This provides an initial guess for the number of infected and then we double this for the number of exposed. We note that these initial conditions are also fine tuned by the ESMDA algorithm. Our population by age group data for each locality comes from the 2018 U.S. Census Bureau estimates. We use the age groupings described in [1], however, this type of data is not available in each racial/ethnic category. As an estimate we take the age grouping data from the specific locality and scale it by the proportion of the population each community represents. This is not completely ideal, but serves as a reasonable estimate. We also note that some population percentages for a given state may add up to more than 100%. This could be due to double counting in some cases. In the end population estimates will have inaccuracies in general and we believe these estimates are sufficient for the current work. We anticipate more accurate information on age groups coming from the 2020 U.S. census when it is made available.

In our analysis, we employ three different inter-population matrices for each of the three intervention periods as in Table 2. In the first period before any mitigation measures, 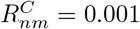 for *n ≠ m*, during the lockdown periods we reduce the off-diagonal terms by an order of magnitude to 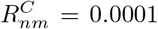, and finally after the lockdown we take the off-diagonal terms to be 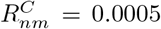, assuming caution in the population. We choose the off-diagonal mixing terms to be fairly small by the following reasoning. Most of the spread of this virus occurs when mask-less, indoors, and with sustained close contact. This implies that, especially during and after the lockdown periods, most of the spread will happen in the home or group settings where precautions are not taken or are infeasible to maintain, an assumption supported by a recent CDC study [22]. This implies that for the majority of the pandemic, members of these specific racial groups are not often having the type of contact conducive to spreading SARS-CoV2 with people outside of their family nor their group of close friends, both of which are likely to consist of members of the same racial/ethnic group. This implies that these groups may in fact evolve somewhat independently of each other with transmissions between racial groups being less common than with in groups. To illustrate, a member of a particular group may become exposed at a place with other groups present, i.e. work or large gathering. This would represent one new infection for this individual and their group. However, they may then spread infection to several immediate or extended family members who likely share race/ethnicity. Thus, one infection across groups can become many infections within the group. This effect would be amplified during times when people are taking precautions when outside of the home.

In practice, when repeating our experiments over a range of values for 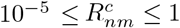 we find that we can best explain the data in the range of 10^*−*4^ to 10^*−*2^ as measured by the *χ*-squared statistic. In addition, the *R*_*t*_(*n*) curves for each group retain the general trend of the initial prior, which is desirable. For values of large mixing (*≥* 10^*−*1^) the groups with the highest number of infections per capita tend to drive the dynamics with the other groups having values for *R*_*t*_(*n*) well below the exponential threshold of *R* = 1. This is unrealistic as it would imply one group is primarily responsible for the majority of transmission which is incompatible with the fact that most of the transmission happens in the home or close group settings.

This effect can be understood through equations (1a) and (1b). Large values for 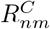 would require a large reduction in *R*_*t*_(*n*) to scale down the transmissions from the group *m* (with a large number of infections) to group *n* (with far fewer infections) so that they are consistent with the number of deaths in the data. Likewise, if 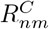 is too small *R*_*t*_(*n*) may be driven up. However, if a particular group is a very small proportion of the population, then an *R*_*t*_(*n*) below *R* = 1 is realistic as interactions with their own groups are less likely and interactions with other groups would be the primary driver of spread. As a result, *R*_*t*_(*n*) would not be driven down just because of a large mixing value. We illustrate some of these differences in Figure 5. We also note that parameters that are well understood, such as the CFR, must change somewhat significantly between groups as well in order to fit the data for large mixing values. For all of the reasons above we keep the off-diagonal mixing values around the order of 10^*−*4^ *−*10^*−*3^.

**Figure 5.**
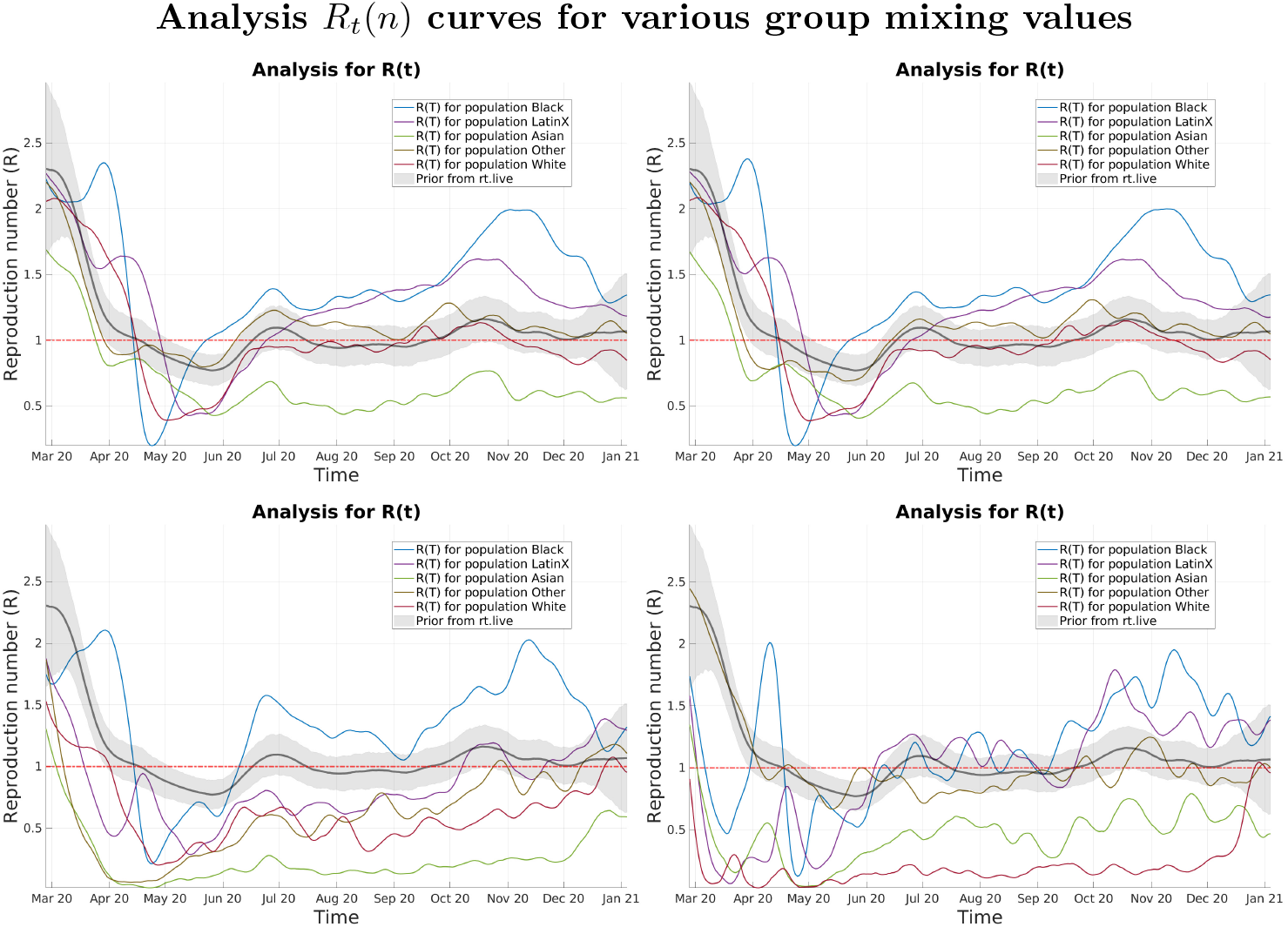
Examples of analysis runs for various values of 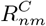 from the District of Columbia (DC). Top: left 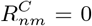, right 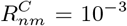. Bottom: left 10^*−*1^, right 6 *×* 10^*−*1^

We performed three types of runs, each designed to study different aspects of possible disparities between groups. In the first run type, we use age contact matrices of all ones (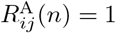 for all age groups *i, j* and populations 1, *…, n*) which would remove any possible effect resulting from differences between the age group contact matrices of the FLCs and the NFLCs. We do this in an effort to detect any possible disparities amongst groups without making prior assumptions of how their contact rates might differ. We further assume the same piece-wise prior for each group on the effective reproductive scaling factor *R*_*t*_(*n*) with large uncertainty (*σ* = 3), allowing the ESMDA smoothing to adjust this based on the data. The prior itself is taken from rt.live [20] for each state that we study. We would expect the recovered *R*_*t*_(*n*) for each group to be somewhat stratified with larger values on average belonging to the FLCs. This is shown in Figure 6 for the District of Columbia where we see a stratification indicating the Black and Latinx communities having the largest values for *R*_*t*_(*n*). We do note the somewhat poor data fit when doing piece-wise updates, this is remedied when allowing for a time-continuous update to *R*_*t*_(*n*) discussed below.

**Figure 6.**
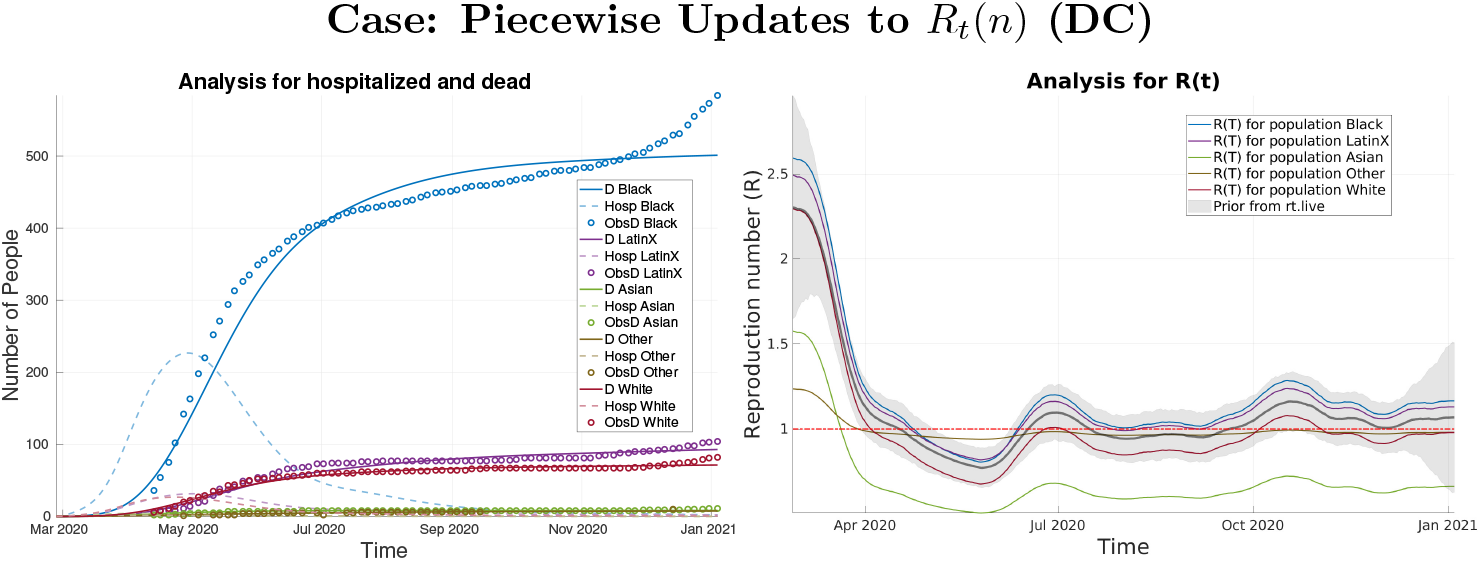
Analysis results for The District of Columbia (DC) with 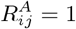 and piece-wise updates to *R*_*t*_(*n*)

In the second run, we repeat the same experiment as above with the exception of using different age contact matrices. Here we use the FLC and NFLC matrices described in Section 4.1. We assign the FLC matrices to subgroups that meet the criteria for disparity as described by the group at covidtracking.com [2], also outlined in Section 1.1. For this second run with FLC and NFLC groups, we typically expect the stratification to be somewhat less dramatic as the increased contact rates among working age and elderly groups in the FLCs can explain increased transmissions and deaths without higher values for *R*_*t*_(*n*), which is again updated as a piece-wise function. An example of these runs is shown in Figure 7 where we see a stratification among groups with the analysis *R*_*t*_(*n*) values typically a bit lower than for the case without age stratification. This is because of the increased contact rates amongst adults in the age-stratified matrices, who are more likely to suffer death, as compared to that of children, who are less likely to suffer death from SARS-Cov2. This means that less spread is needed to account for the number of deaths in the data. We also note that the Black community *R*_*t*_(*n*) curve now lies atop the Latinx curve having come down more from the previous run. This is because the Black community met the criteria outlined in Section 1.1 to be an FLC while the Latinx community did not in the District of Columbia. As a result, the Black community was assigned the FLC matrix which has higher contact rates for working-age and older generation age individuals, again reducing the amount of spread needed to explain deaths.

**Figure 7.**
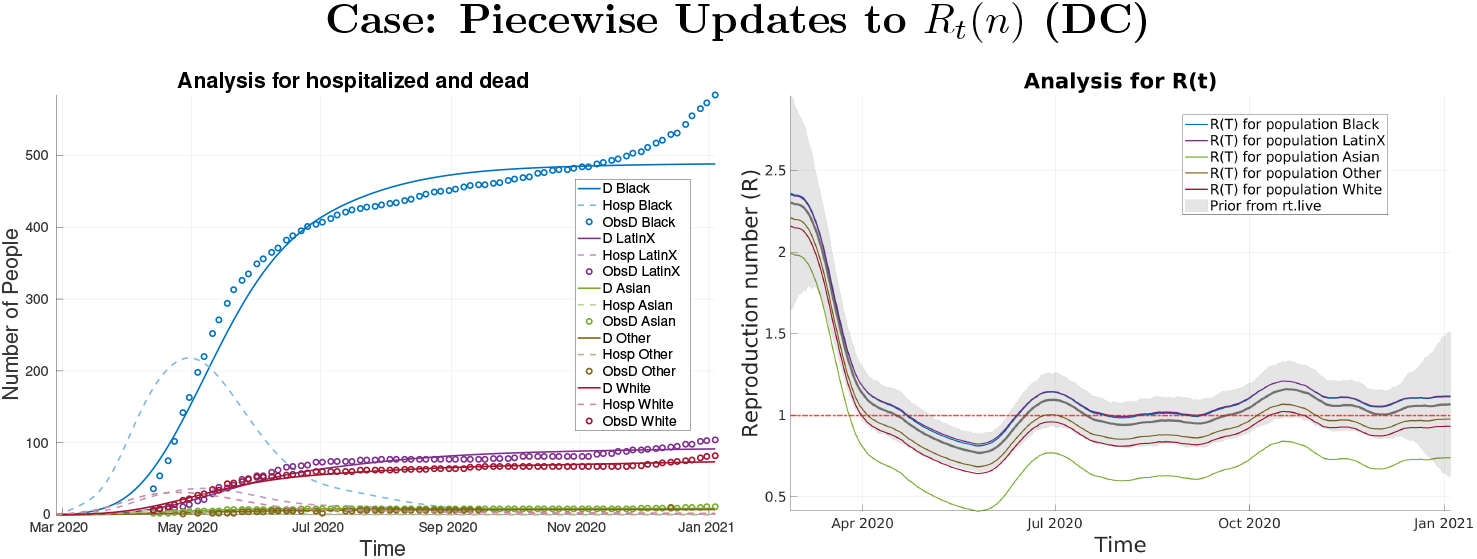
Analysis results for The District of Columbia (DC) with age stratified *R*^*A*^ and piece-wise updates to *R*_*t*_(*n*)

This effect can also be understood through equations (1a) and (1b), specifically the product of 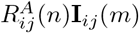 when *n* = *m*. If contact rates for working-age and older individuals are higher, the number of infections attributed to those groups is increased. As a result, the scaling factor *R*_*t*_(*n*) may need to be reduced if those infections are overestimated and inconsistent with the number of deaths in the data. The specific contact rates are difficult to determine and a study to determine them is encouraged.

Finally, we complete a third run where we allow for a time-continuous estimation of *R*_*t*_(*n*) using a decorrelation length of 10 days, the CDC recommended time that an individual should quarantine after exposure [23]. Here the priors for *R*_*t*_(*n*) used for each group are taken directly from the analysis *R*_*t*_(*n*) that comes from the second run. We decrease the uncertainty to *σ* = 0.5 and employ the FLC and NFLC matrices in the same way as previously described in Section 4.1.

For all run types, we have three intervention periods where we begin with large contact rates at the beginning of the pandemic, severely reduced contact rates during a given state’s mitigation (lockdown) period, and slightly reduced contact rates after a given state re-opening date. How we scale these rates for each age group is discussed in Section 4.1 and given in Table 1. The dates, in addition to the starting date of each simulation, for each of the states we study can be found in Table 2. The start date is chosen to be the first day given by our prior which comes from rt.live. The date of the first intervention is chosen when a state began to close schools and the beginning of the introduction of the White House Coronavirus Task Force’s initial “15 days to slow the spread” campaign which prompted many Americans to take precautions ahead of their individual states mandates.

#### 4.2.1 The state of Alaska (AK)

The groups we consider for Alaska are shown in Table 3. Immediately striking is a major disparity revealed in the number of deaths between the White and American Indian and Alaskan Native (AIAN) communities. The White population makes up about 65% of the total population of the state while the AIAN population is only about 15%, yet they have comparable numbers of deaths. This can be seen in the analysis for hospitalized and dead plots in Figure 8. This disparity may be due to the large proportion of the AIAN community which lives far north of the Arctic Circle in regions where it is desirable to be indoors (where transmission is more likely) most of the year, particularly in the winter months. There is also far less medical infrastructure in these regions which can contribute to higher death rates. There is also a noticeable disparity amongst the Native Hawaiian and Pacific Islander (NHPI) community which makes up only about 1% of the population. In the analysis of exposed and infected plots of Figure 8, the number of infections of this group is above both that of the Asian and Black communities which make up 6% and 3% of the population respectively. In the same three figures, we also see that the number of total infections–as estimated by the analysis up to the last data point on January 3, 2020–for the NHPI population is higher than that of the Black population. We also see evidence of these disparities in the analysis of the *R*_*t*_(*n*) functions for all three types of runs. In the ensemble of *R*_*t*_(*n*) plots for the first two types of runs with piece-wise updates and large uncertainty, we see that in Figure 18 (Appendix B) stratification of the curves occurs with the most affected groups above the least effected groups for the majority of the integration interval. In the first scenario, where all entries of the age contract matrices are equal to one, we observe slightly higher values for *R*_*t*_(*n*) than in the second case where we employ the age-based contact matrices described in Section 4.1. This is due to the higher contact rates in the contact matrices for more susceptible age groups (adults) in the second case. Otherwise, *R*_*t*_(*n*) must be larger to account for the rate of spread. It is also notable that in the first two cases the rapid increase in death during the winter surge is somewhat missed by the analysis.

**Figure 8.**
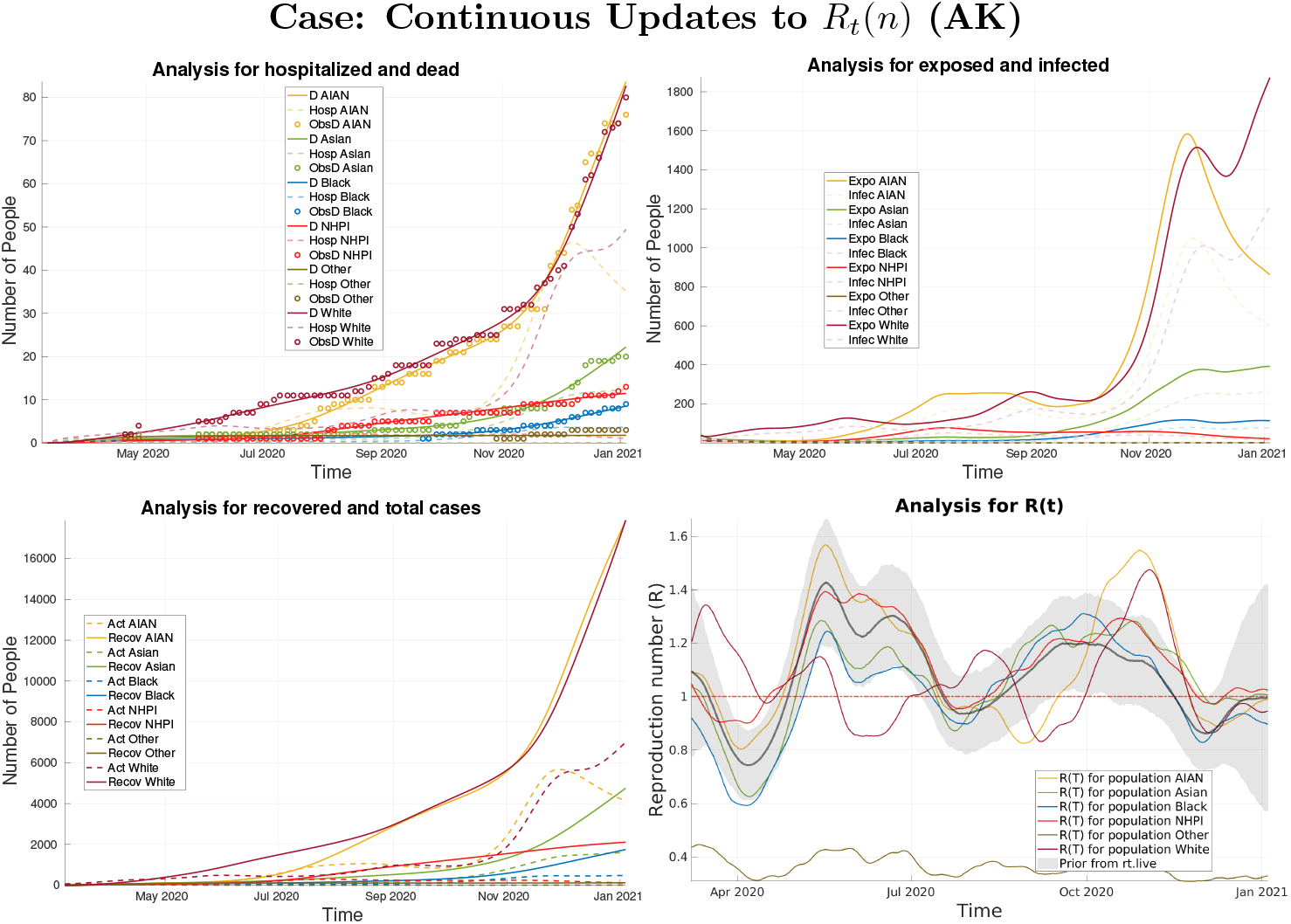
Analysis results for the continuous update case for the state of AK.

**Figure 9.**
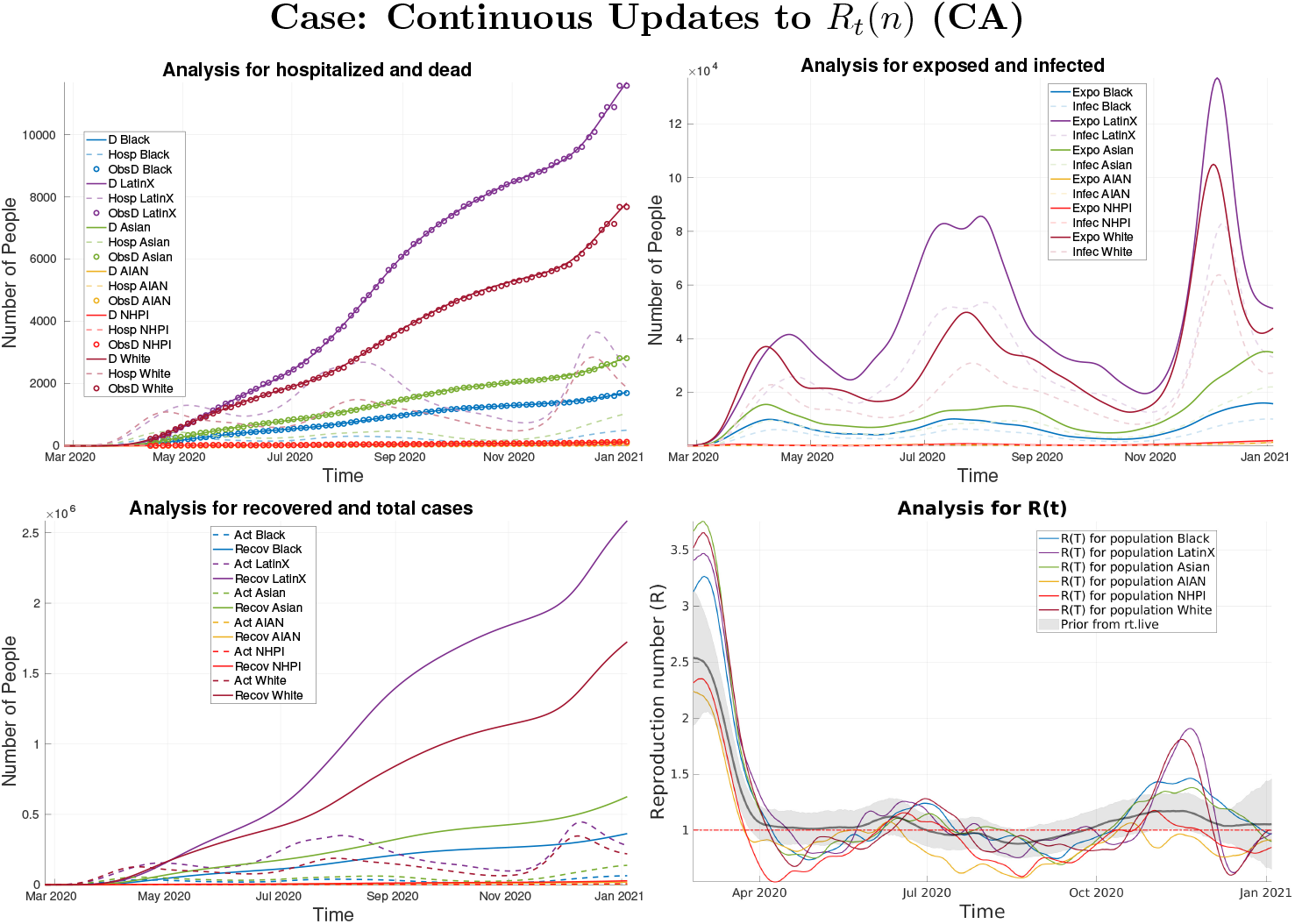
Analysis results for the continuous update case for the state of CA.

**Figure 10.**
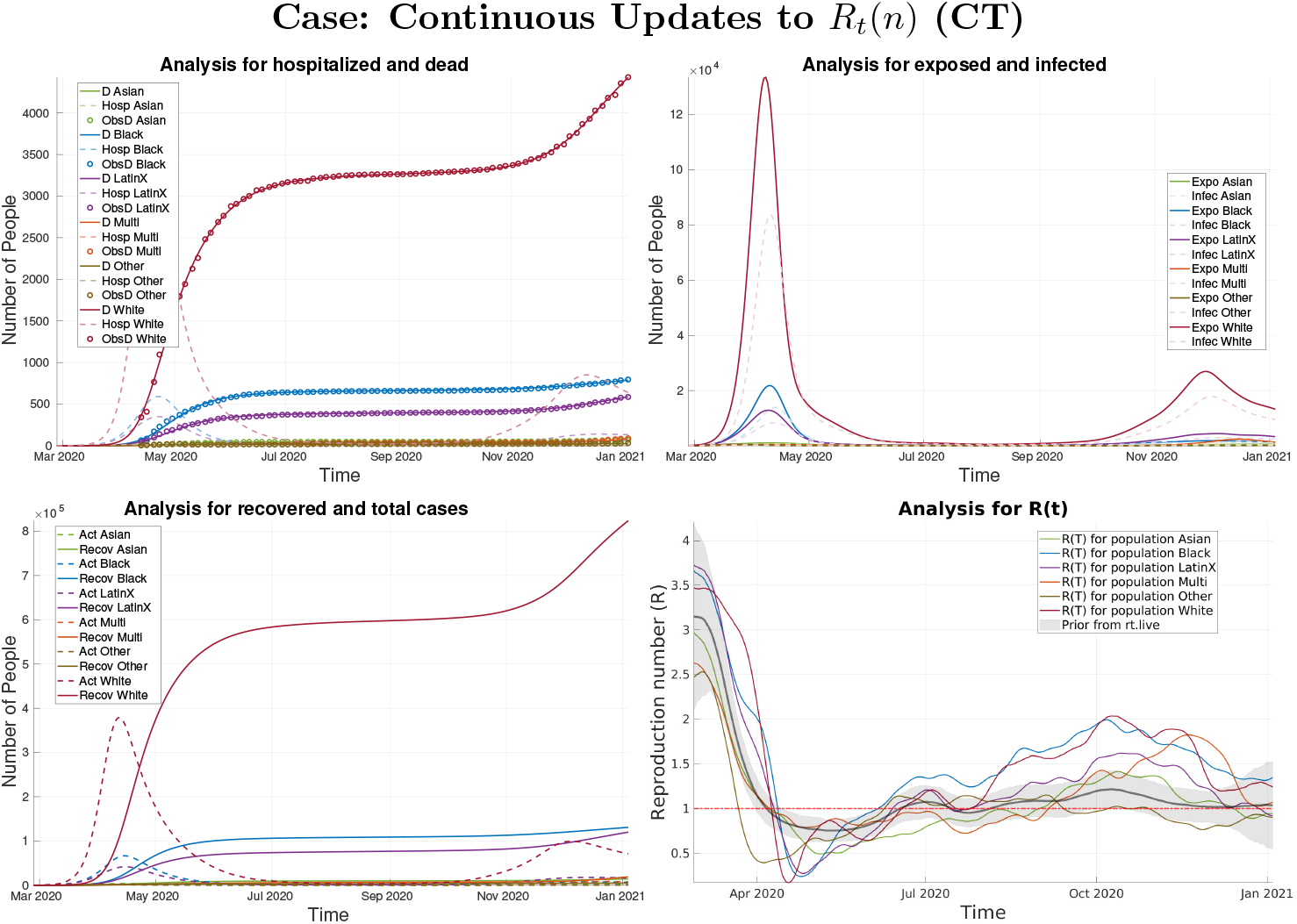
Analysis results for the continuous update case for the state of CT.

**Figure 11.**
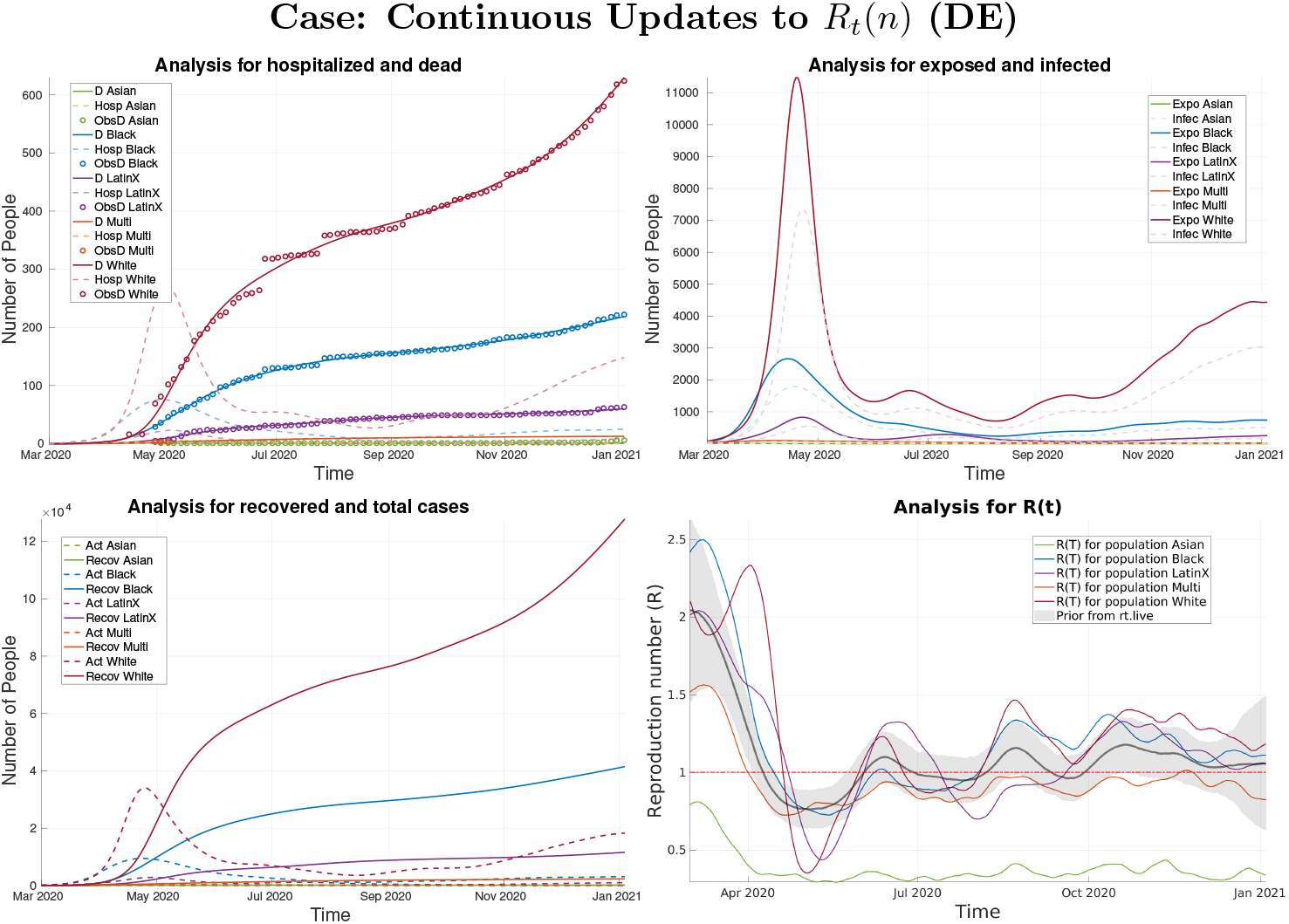
Analysis results for the continuous update case for the state of DE.

**Figure 12.**
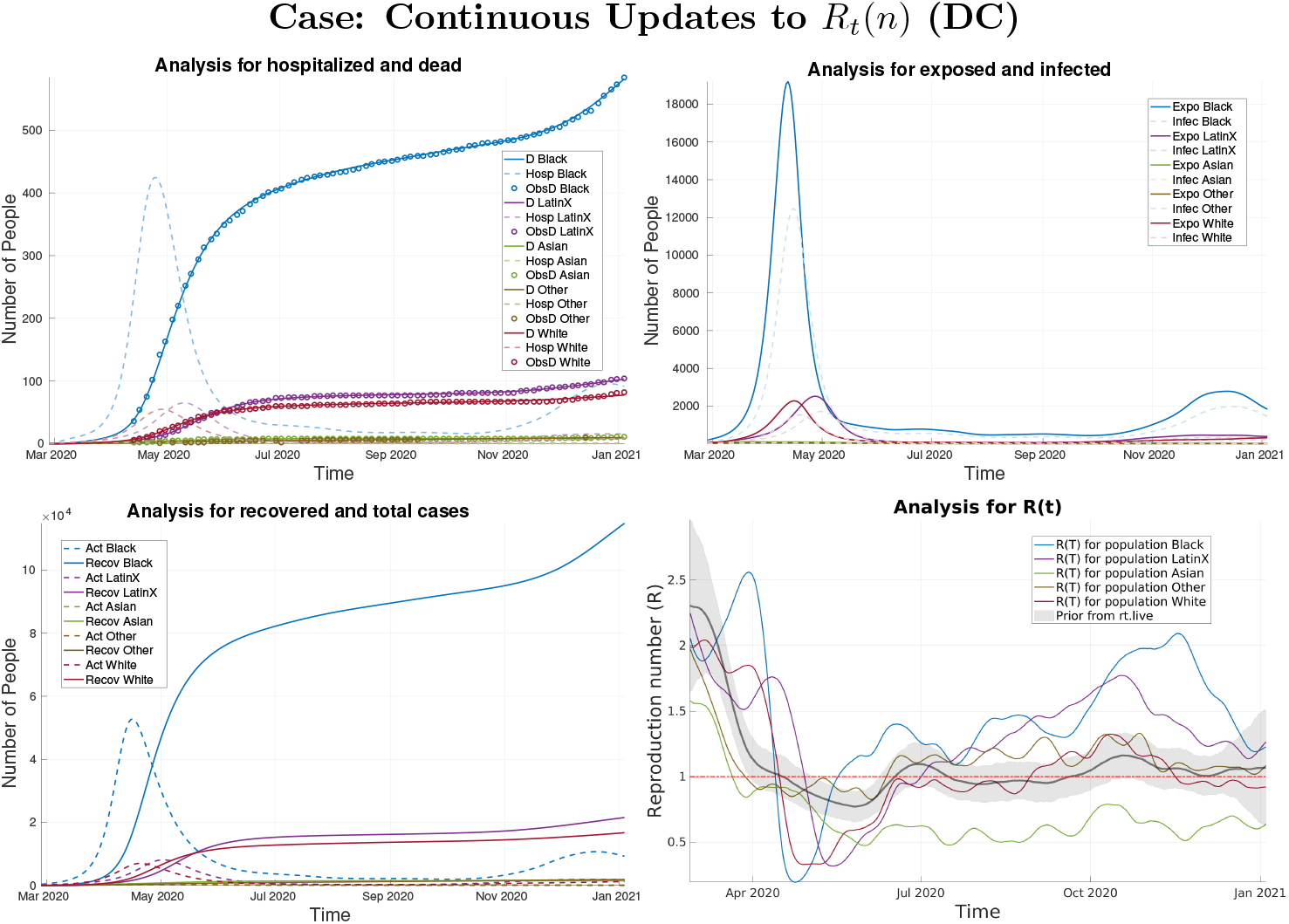
Analysis results for the continuous update case for the District of Columbia.

**Figure 13.**
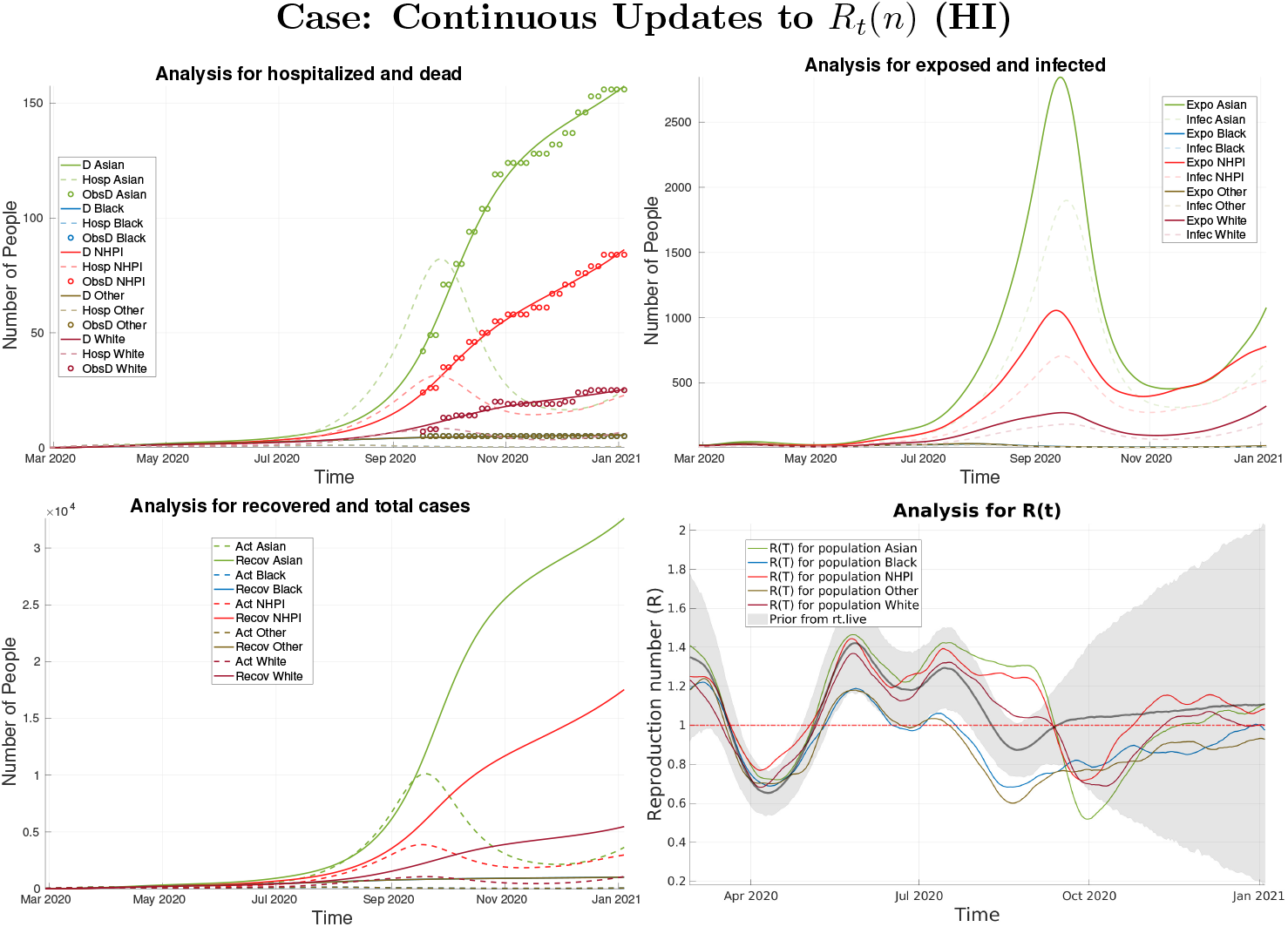
Analysis results for the continuous update case for the state of HI.

**Figure 14.**
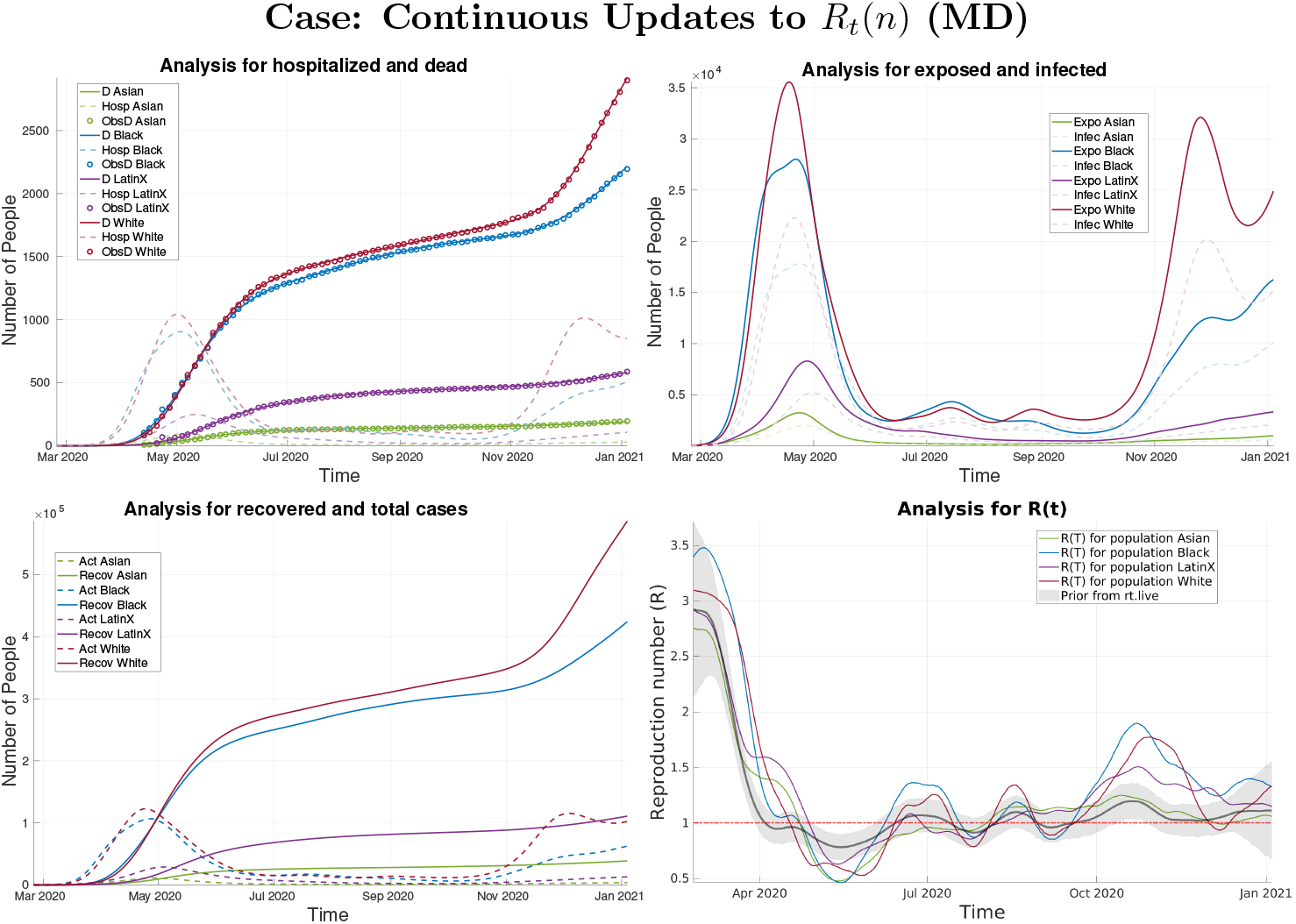
Analysis results for the continuous update case for the state of MD.

**Figure 15.**
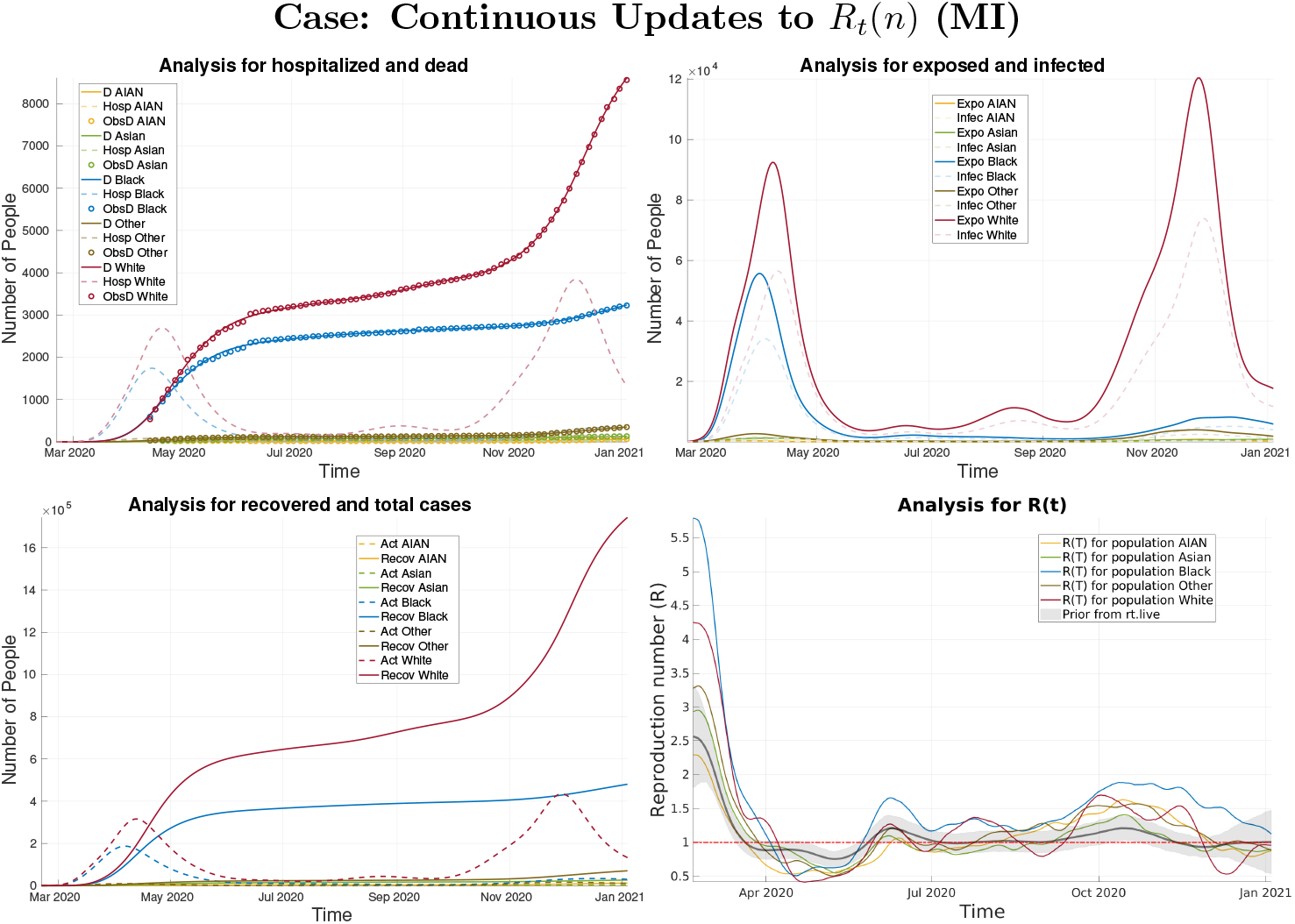
Analysis results for the continuous update case for the state of MI.

**Figure 16.**
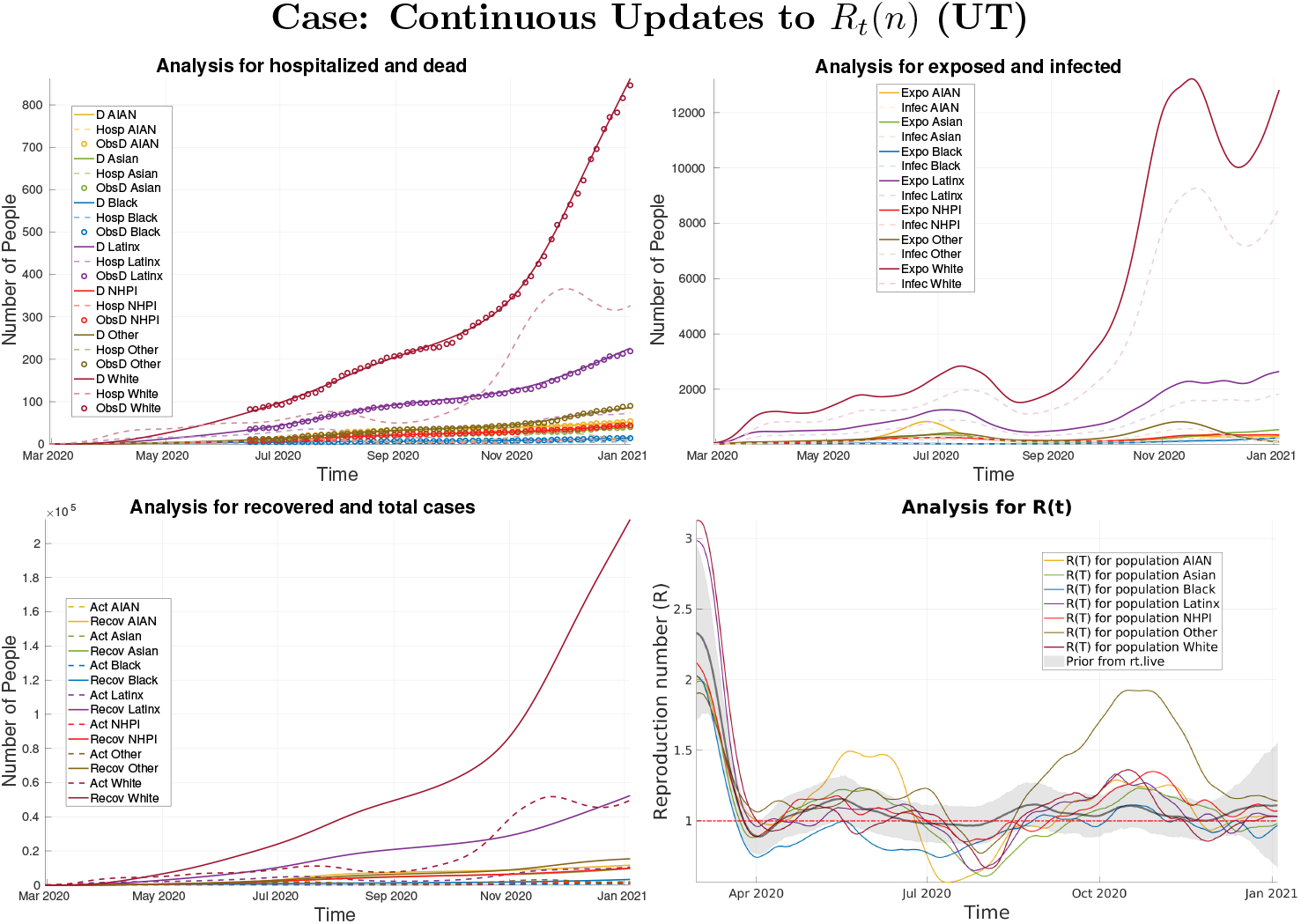
Analysis results for the continuous update case for the state of UT.

**Figure 17.**
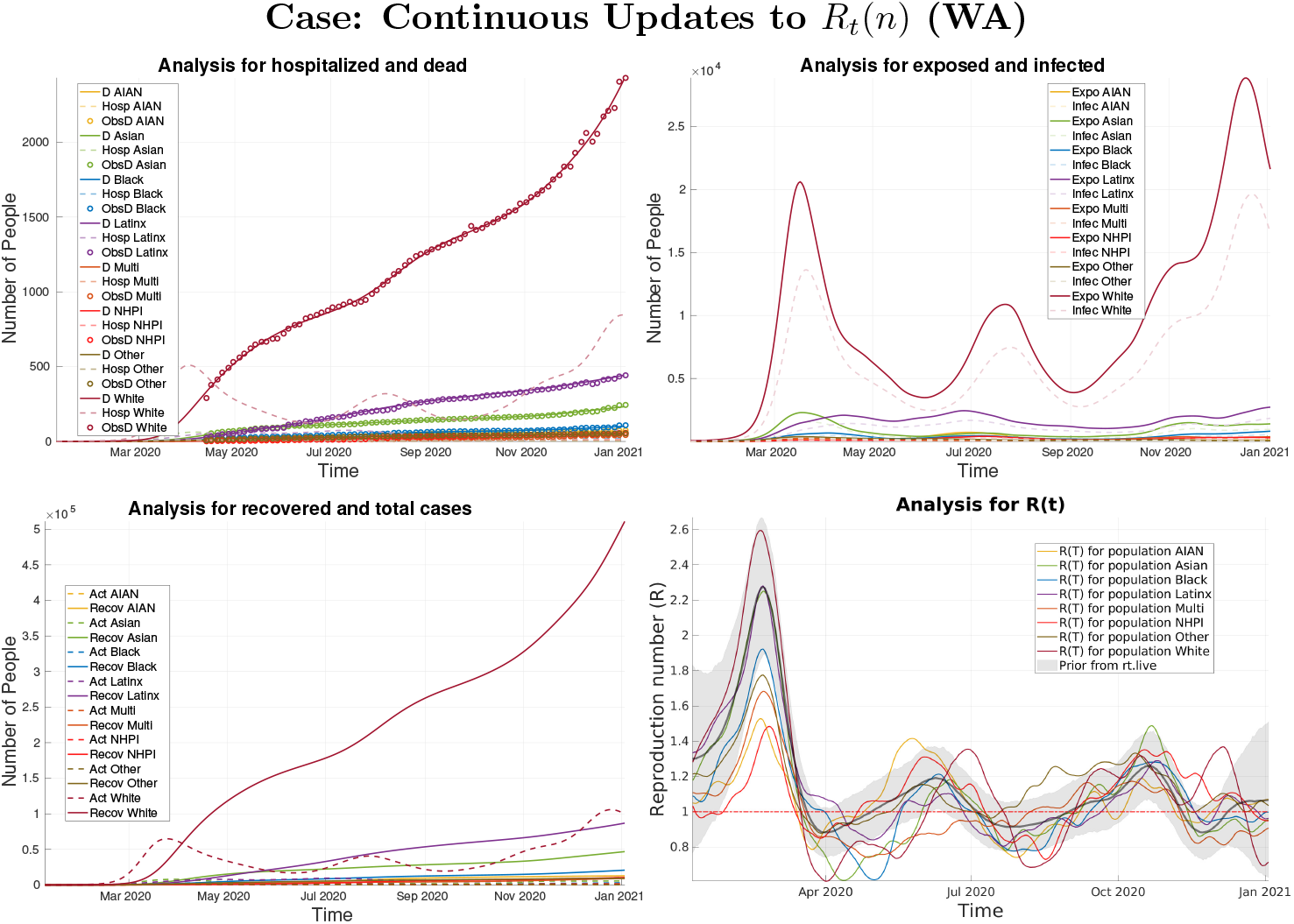
Analysis results for the continuous update case for the state of WA.

**Figure 18.**
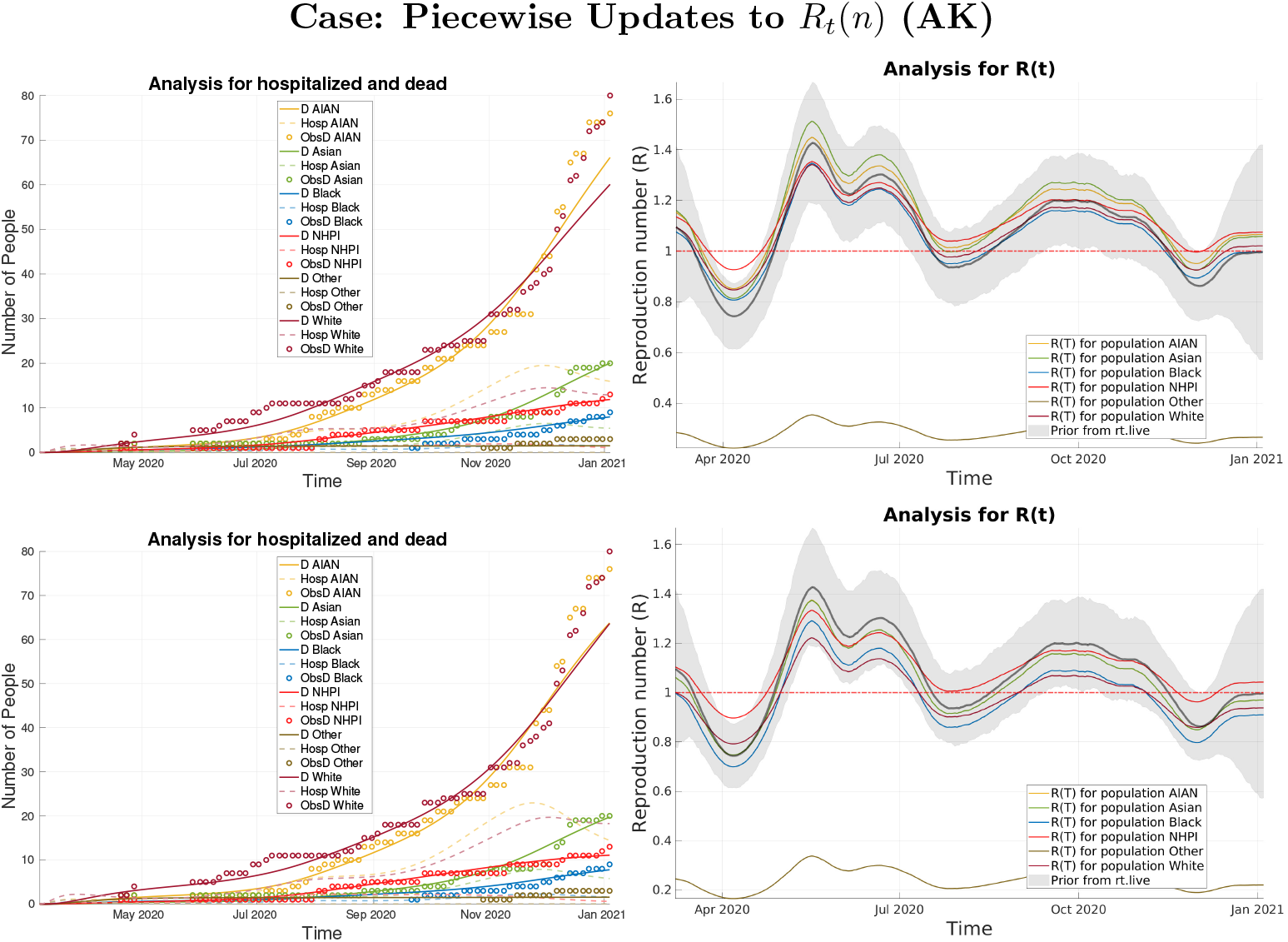
Analysis results when using peicewise updates to *R*(*t*) for AK. Top row: *R*^*A*^ with entries of all ones. Bottom Row: *R*^*A*^ for FLCs and NFLCs.

In the most realistic case, where we allow for a continuous update of *R*_*t*_(*n*) with a 10-day decorrelation length, this winter surge is captured and we see a far richer difference in the analysis *R*_*t*_(*n*)’s between groups. For the White population, we notice a significantly higher *R*_*t*_(*n*) from the start of the pandemic through mid-April, this suggests that this was the first group affected, likely because the virus would have arrived in larger cities, such as Anchorage, where a large proportion of the state’s White population resides. After that time we see *R*_*t*_(*n*) typically below *R* = 1 while spread becomes exponential for most of the other racial/ethnic groups once the virus reaches beyond cities, into the regions with larger AIAN populations. With the exception of the beginning of the pandemic, we also note that *R*_*t*_(*n*) dips further below *R* = 1 and more often for the White population. Notably, *R*_*t*_(*n*) remains well below *R* = 1 for the “Other” group, while Other represents 8% of the population. The reason for this may be that many deaths actually in this group are yet unreported or were reported as members of one of the other groups the state of Alaska considers.

#### 4.2.2 The state of California (CA)

The groups we consider for California are shown in Table 3. The Latinx and White groups make up 39% and 37% of the population of California respectively. As can be seen in the analysis for hospitalized and dead plots, both groups in Figures 19 (Appendix B) and 9 evolve very similarly early on but begin to diverge from each other after the first set of restrictions are lifted at the end of May 2020. The Latinx population begins to overtake the White population in deaths and infections for all three run types with a continually widening gap. The Latinx community is the majority in the state of California, however, only by about 3%. The disparity between infections and deaths after the lifting of restrictions may be related to which type of employment is more common in each group. According to the U.S. Equal Employment Opportunity Commission, White/Asian individuals comprise about 87% and 90% of the high tech jobs in the San Francisco-Oakland-Fremont and Santa Clara County regions, respectively, while Black/Hispanic individuals comprise only about 10% and 8% of the high tech jobs in each of those regions [24]. In March 2020, out of the roughly 19.17 million individuals that make up the labor force in California [25], about 1.87 million of those jobs were in the technology field [26]. That is, almost 1 in every 10 working-aged individuals in California would have a job considered in the technology sector of the economy. Since such jobs are more easily amicable to being conducted remotely, and because of the large racial disparity we see for the demographics of such jobs, this could be one influencing factor in the disproportionate spread of COVID-19 in the Latinx community.

**Figure 19.**
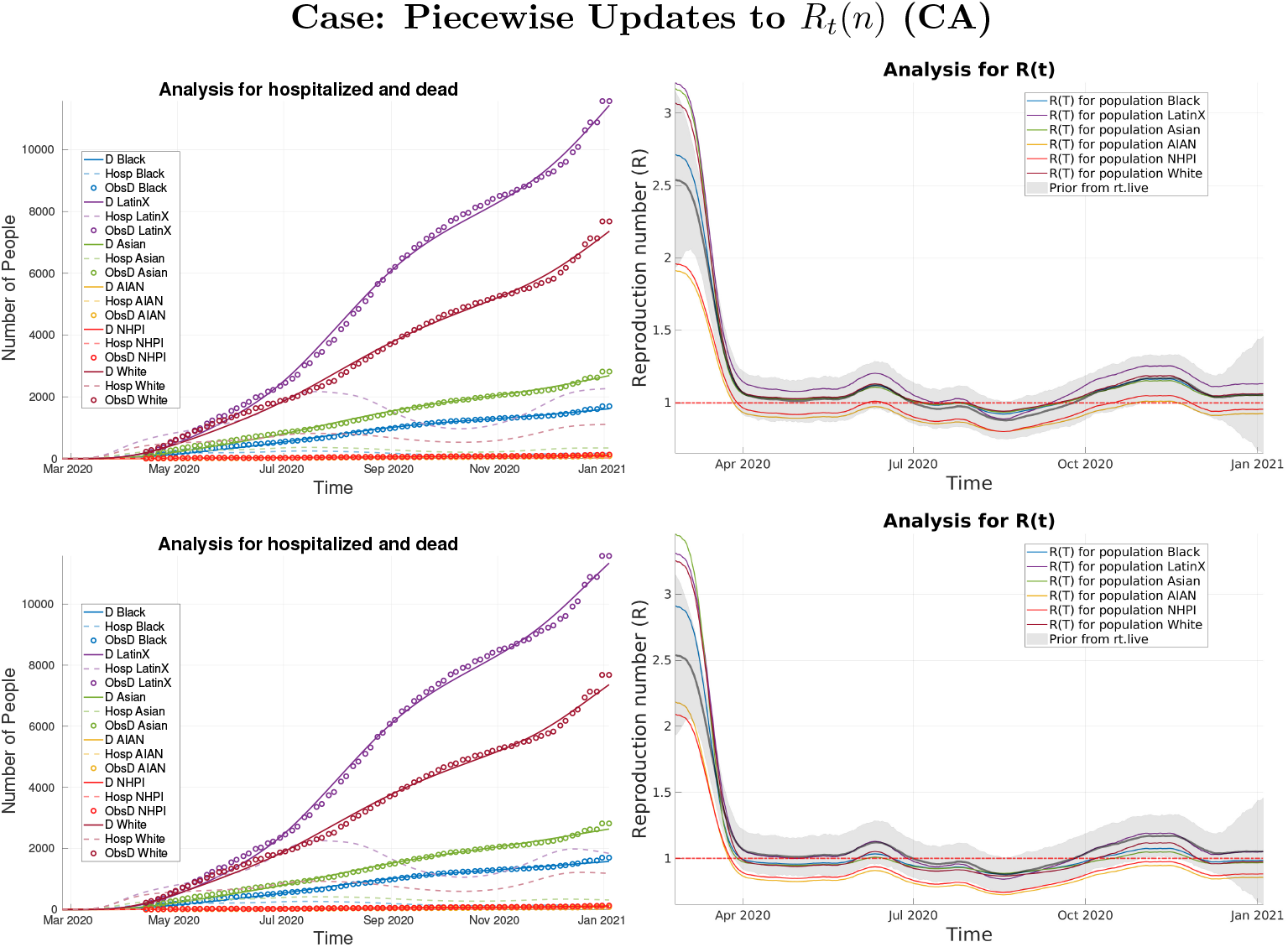
Analysis results when using peicewise updates to *R*(*t*) for CA. Top row: *R*^*A*^ with entries of all ones. Bottom Row: *R*^*A*^ for FLCs and NFLCs.

In the case of California, the Latinx community met the criteria to be classified as an FLC as described in Section 1.1. When we examine the analysis *R*_*t*_(*n*) curves for the case of a contact matrix of all ones (Figure 19, top, Appendix B), we do see some stratification occurring among groups with the Latinx community a bit above the others. However, when employing the age-based contact matrices (Figure 19, bottom, Appendix B) this stratification disappears between the two majority communities. In the case of a time-continuous update to the *R*_*t*_(*n*) curve (Figure 9), the curve for the Latinx community is, on average, above or similar to the White Group.

#### 4.2.3 The state of Connecticut (CT)

The groups we consider for Connecticut are shown in Table 3. In the state of Connecticut, the Latinx and Multi-Racial populations meet the criteria for the disparity in the disproportionate number of confirmed cases as outlined in Section 4.2. The Latinx community makes up about 16% of the state’s population, while the Black and White communities make up 10% and 67%, respectively. All other groups considered in this state’s analysis make up less than 5% of the population, with the Multi-Racial group making up about 2%. According to the case count data, 26% of confirmed cases come from the Latinx community while only 11% come from the Black community. However the Black community has more deaths, and for all three run types, they exhibit more overall cases. It is important to again note that confirmed cases may not be representative of all the actual cases, which is the reason why we do not assimilate that data. It may be possible that one community has more access to testing than another or maybe more likely to get tested in general. The fact that the Black community makes up 6% less of the population of Connecticut, yet accounts for 3% more of the deaths (at the time of writing) may be an indication of disparities. The low number of confirmed cases compared to the Latinx community suggests either a higher case fatality rate (CFR) or poor access to testing for the Black community. The CFR for each group is estimated in our ESMDA process, however, we do not detect any appreciable difference between these groups in this state for any of the run types.

When we examine the analysis *R*_*t*_(*n*) functions using piece-wise priors (Figure 20, Appendix B), we see that during the first intervention period, *R*_*t*_(*n*) is much lower for the White population than all of the others, possibly relating to the types of jobs held by the groups, their access to C.A.R.E.S. act aid, or reserve funds to fall back on. This apparent difference is lessened somewhat in the time-continuous case. In Figure 10 we do see the effect of the first intervention period on *R*_*t*_(*n*) between April and May with a sharper dip for all groups with a minimum value amongst the White population. After the reopening, we see an increase in *R*_*t*_(*n*) for all groups with the White and Black populations sustaining the highest, and comparable, values for *R*_*t*_(*n*). The Latinx community maintains *R*_*t*_(*n*) values less that of the White and Black populations even though they represent a larger proportion of the population than that of the Black community. This again suggests the detection of some disparity for this Black population.

**Figure 20.**
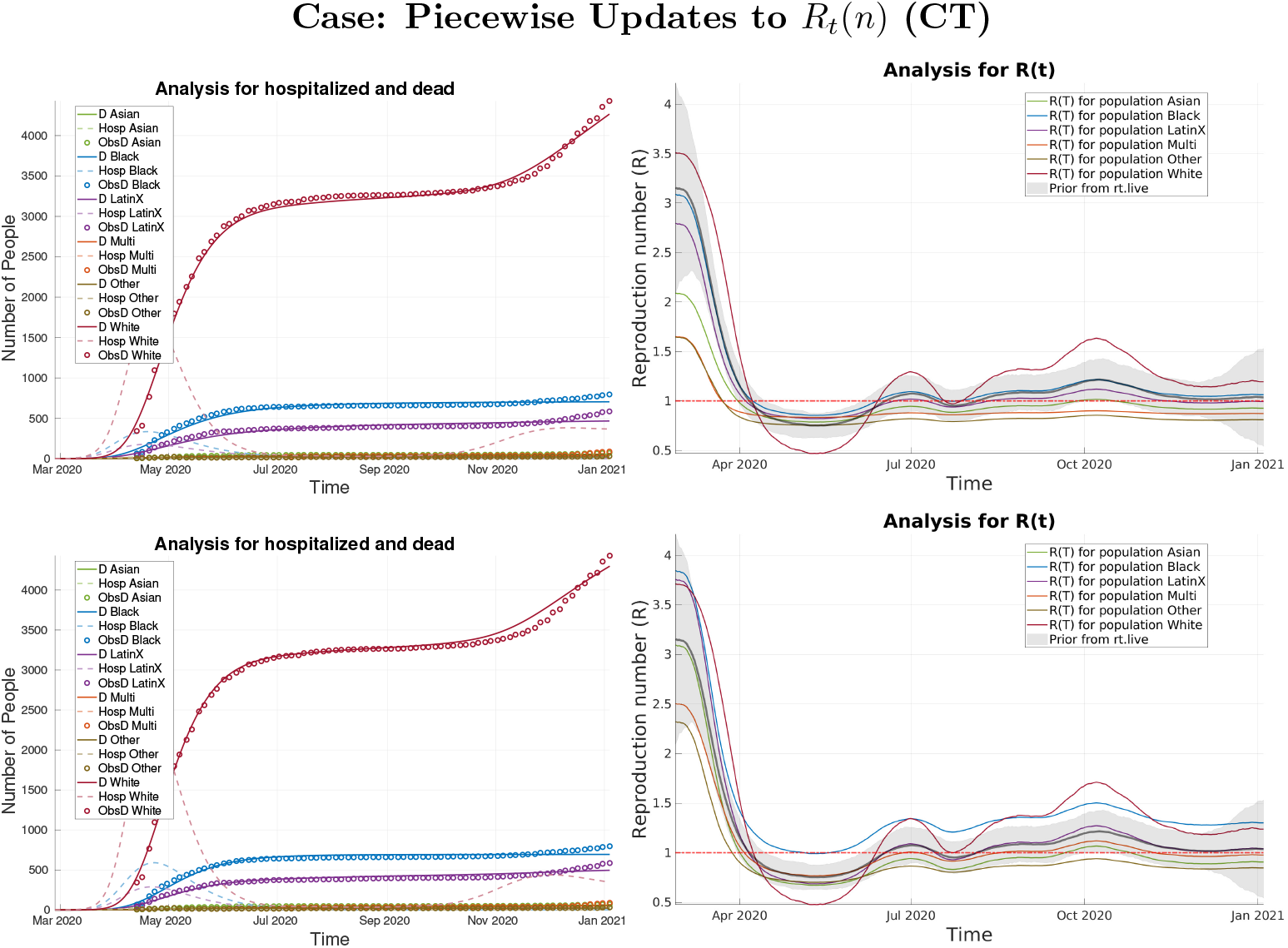
Analysis results when using peicewise updates to *R*(*t*) for CT. Top row: *R*^*A*^ with entries of all ones. Bottom Row: *R*^*A*^ for FLCs and NFLCs.

#### 4.2.4 The state of Delaware (DE)

The groups we consider for Delaware are shown in Table 3. The Latinx community fits the criteria described in Section 4.2 for an FLC, making up about 9% of the population but accounting for 18% of confirmed cases. When examining the analysis *R*_*t*_(*n*) values for the case with a contact matrix of all ones, in Figure 21 we see a stratification of *R*_*t*_(*n*) amongst the groups. During the intervention period, we find that the White population has the lowest values of *R*_*t*_(*n*) of the three most populous groups in the state with the Latinx population having the highest. The very low value for the Asian community is notable given that they are 4% of the population, however, the state of Delaware includes the NHPI population in this group and some deaths may be reported in the Other or Multi groups. When using the age-based contact matrices and piece-wise updates to *R*_*t*_(*n*), with the Latinx community as an FLC, this stratification is lessened during the intervention period (Figure 21, Appendix B). This deeper dip in *R*_*t*_(*n*) for the White Group during the intervention period is also present in the continuous update case shown in Figure 11. Interestingly, the Black community maintains a much higher value for *R*_*t*_(*n*) during this same time period suggesting that they may also be better classified as a FLC in this state even though they are not flagged as being such.

**Figure 21.**
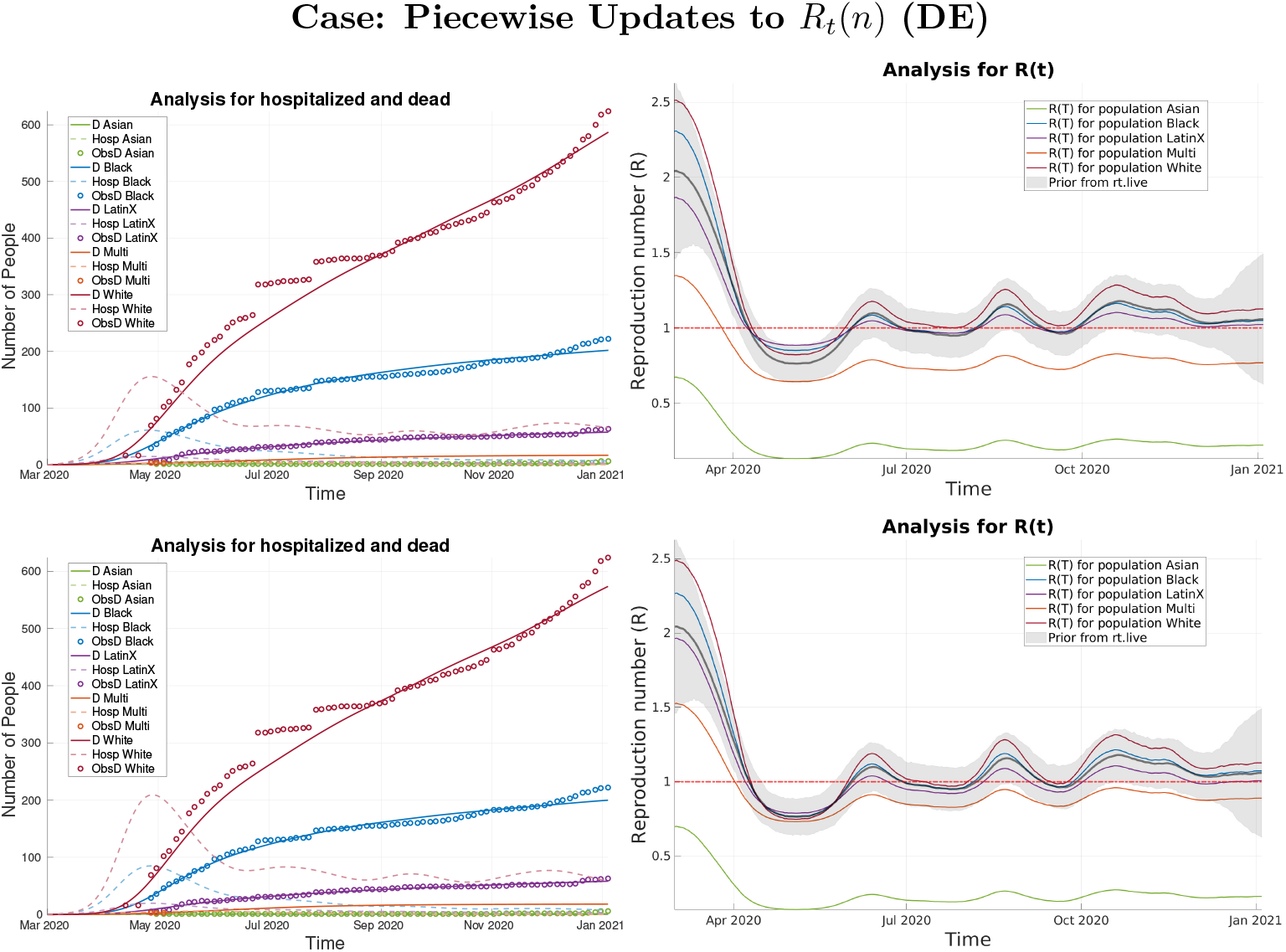
Analysis results when using peicewise updates to *R*(*t*) for DE. Top row: *R*^*A*^ with entries of all ones. Bottom Row: *R*^*A*^ for FLCs and NFLCs.

#### 4.2.5 The District of Columbia (DC)

The groups we consider for The District of Columbia are shown in Table 3. In the District of Columbia, the Black and Latinx populations meet the criteria of an FLC. The Black community comprises 46% of the population and represents 75% of the deaths, while the Latinx community comprises about 11% of the population and represents 25% of confirmed cases and 13% of deaths. In stark contrast, the White population comprises 41% of the population and makes up only 10% of the deaths. In the cases with piece-wise updates to *R*_*t*_(*n*), we see in Figure 22 (Appendix B) that the stratification of the *R*_*t*_(*n*) curves with our FLCs are consistently above those for the NFLCs. We note that the Black and Latinx populations have similar analysis *R*_*t*_(*n*) curves for these runs, however, the Black community has a much more substantial population total. In terms of the number of active cases and total cases, we see in the relevant analysis plots that the Latinx community overtakes the White population despite being a much smaller proportion of the population. A possible disparity unique to the Latinx community may be evident in the analysis *R*_*t*_(*n*) curves for the continuous-time update runs shown in Figure 12 where we see the Black and White population’s *R*_*t*_(*n*) values well below that of the Latinx community’s during the intervention period. These disparities may be related to a group’s ability to remain in “lockdown” by receiving stimulus money or boosted unemployment coming from the C.A.R.E.S. act. In many cases, undocumented workers were unable to receive this federal assistance, hindering one’s ability to remain home possibly contributing to increased rates of spread. After the reopening, we see both the Latinx and Black population’s *R*_*t*_(*n*) curves go above the exponential spread threshold of *R* = 1, while the White populations remain below this threshold until September, where it then moves slightly above that threshold. In the absence of federal financial assistance, groups that have more individuals able to work from home will likely have less spread.

**Figure 22.**
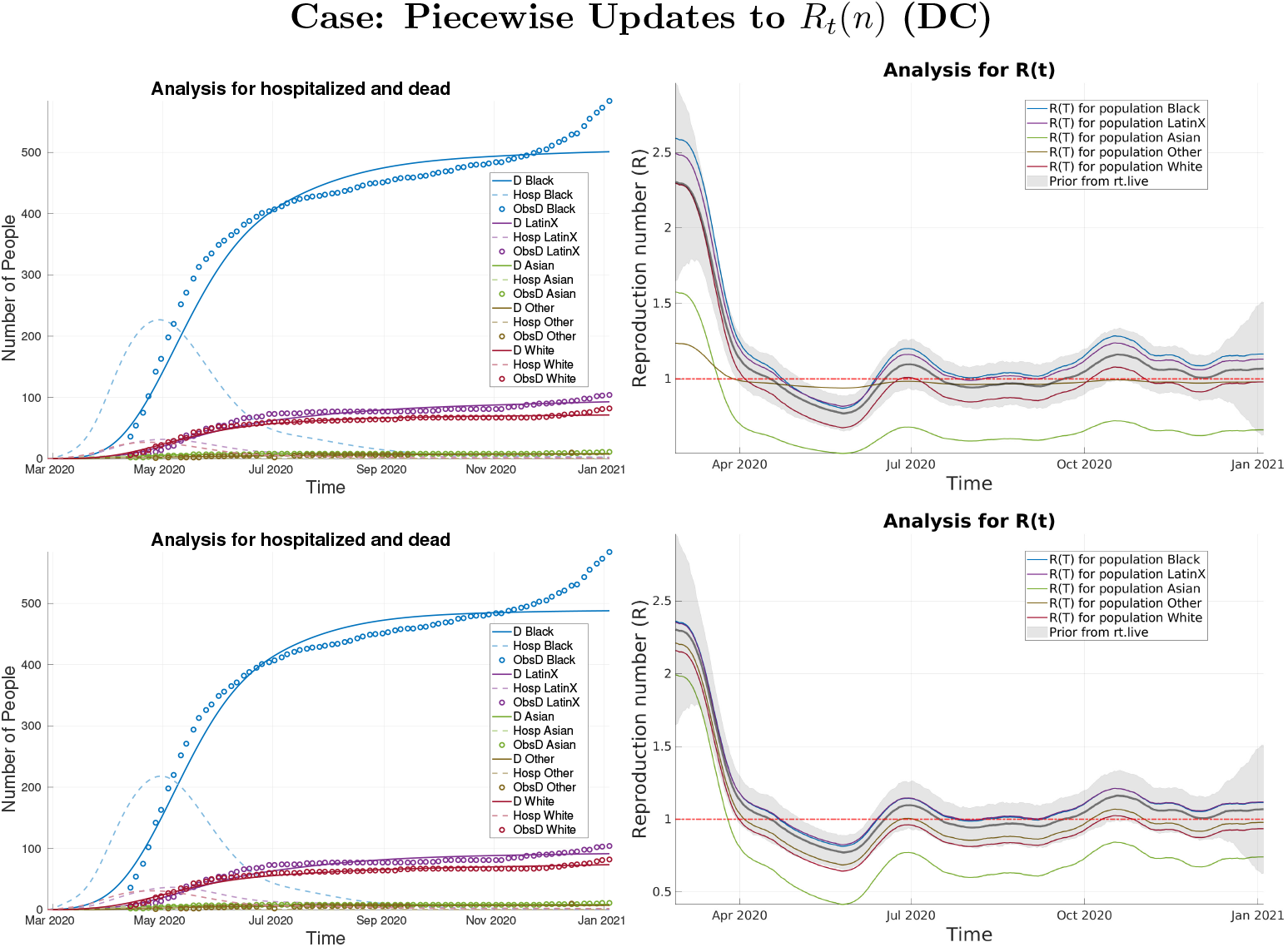
Analysis results when using peicewise updates to *R*(*t*) for DC. Top row: *R*^*A*^ with entries of all ones. Bottom Row: *R*^*A*^ for FLCs and NFLCs.

#### 4.2.6 The state of Hawaii (HI)

The groups we consider for Hawaii are shown in Table 3. The Asian and NHPI populations meet the FLC criteria outlined in Section 4.2. The Asian community makes up 38% of the population and 57% of the deaths, while the NHPI community makes up 10% of the population with 43% of confirmed cases and 31% of the deaths. In the run with contact matrices of all ones, we see in Figure 23 (Appendix B) that there is a stratification in the analysis *R*_*t*_(*n*) with the Asian and NHPI populations maintaining higher values of *R* over all the other groups. The stratification persists when using the age-based contact matrices with the piece-wise updates to *R*_*t*_(*n*), as shown in the bottom row of Figure 23. When examining the analysis *R*_*t*_(*n*) for the time-continuous update case in Figure 13, the stratification during the initial intervention period is reduced but still present with the NHPI community having the largest values of *R*_*t*_(*n*). After the reopening, the stratification persists and increases into the summer months with the Asian population having the largest values of *R* followed by the NHPI population. After September, *R*_*t*_(*n*) is below *R* = 1 for all groups through the end of October before surging again. In this period, the Asian community has the lowest values of *R*_*t*_(*n*) possibly due to the period before where they had the largest, causing enough infections to lower the number of susceptible individuals in the group.

**Figure 23.**
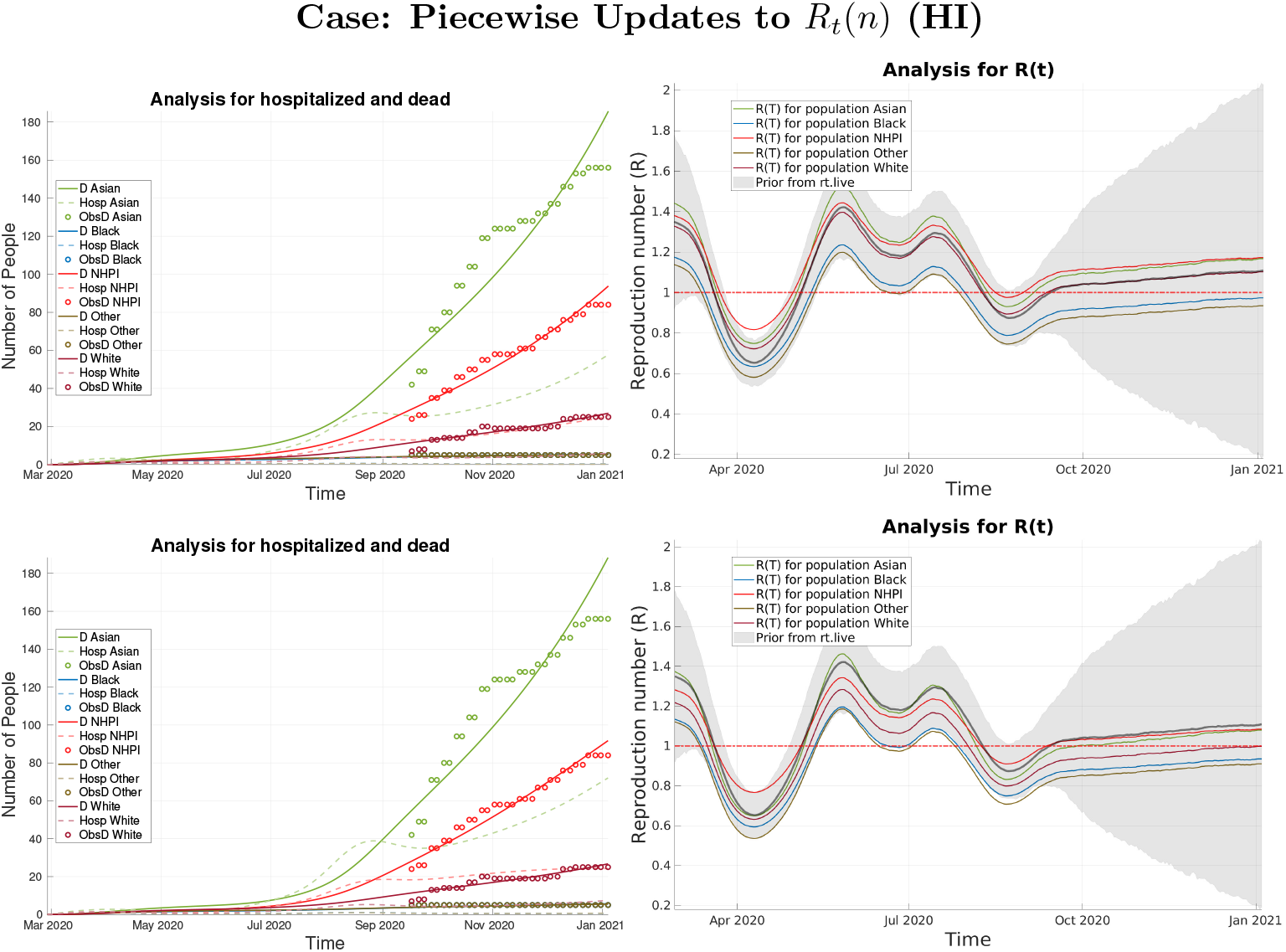
Analysis results when using peicewise updates to *R*(*t*) for HI. Top row: *R*^*A*^ with entries of all ones. Bottom Row: *R*^*A*^ for FLCs and NFLCs.

#### 4.2.7 The state of Maryland (MD)

The groups we consider for Maryland are shown in Table 3. For the state of Maryland, the Latinx community meets the threshold for an FLC as outlined in Section 4.2, as they comprise 10% of the population yet account for 20% of the confirmed cases. Indeed, for all three run types, we see that the Latinx community has the highest values of the analysis *R*_*t*_(*n*) during the intervention period, compared to the other populations observable in Figures 24 (Appendix B), and 14. After the intervention period, we do see an almost steady spread around *R* = 1 for this group when piece-wise updates to *R*_*t*_(*n*) are used (Figure 24). In the case of continuous-time updates to *R*_*t*_(*n*), we see evidence of a fall surge beginning in October for all populations shown in Figure 14. While the Black community did not meet the threshold for an FLC, we see evidence that they could still be one according to our runs and analysis *R*_*t*_(*n*). Comprising 29% of the population of the state, they suffer 37% of the deaths compared to that of the White Group which makes up 51% of the population and 49% of the deaths. Indeed, in the analysis for the exposed and infected plots in Figure 14, we see that before April and between May and September, the Black and White populations have roughly the same number of infections and exposures despite the differences in percent make up of the total population. This suggests a disparity that may be tied to the types of work that the majority of people in each group comprise, such as stay-at-home work versus working at a place of high exposure. It may also be that the case fatality rate is higher for one group than the other and there were fewer infections in the Black community than predicted by the assimilation.

**Figure 24.**
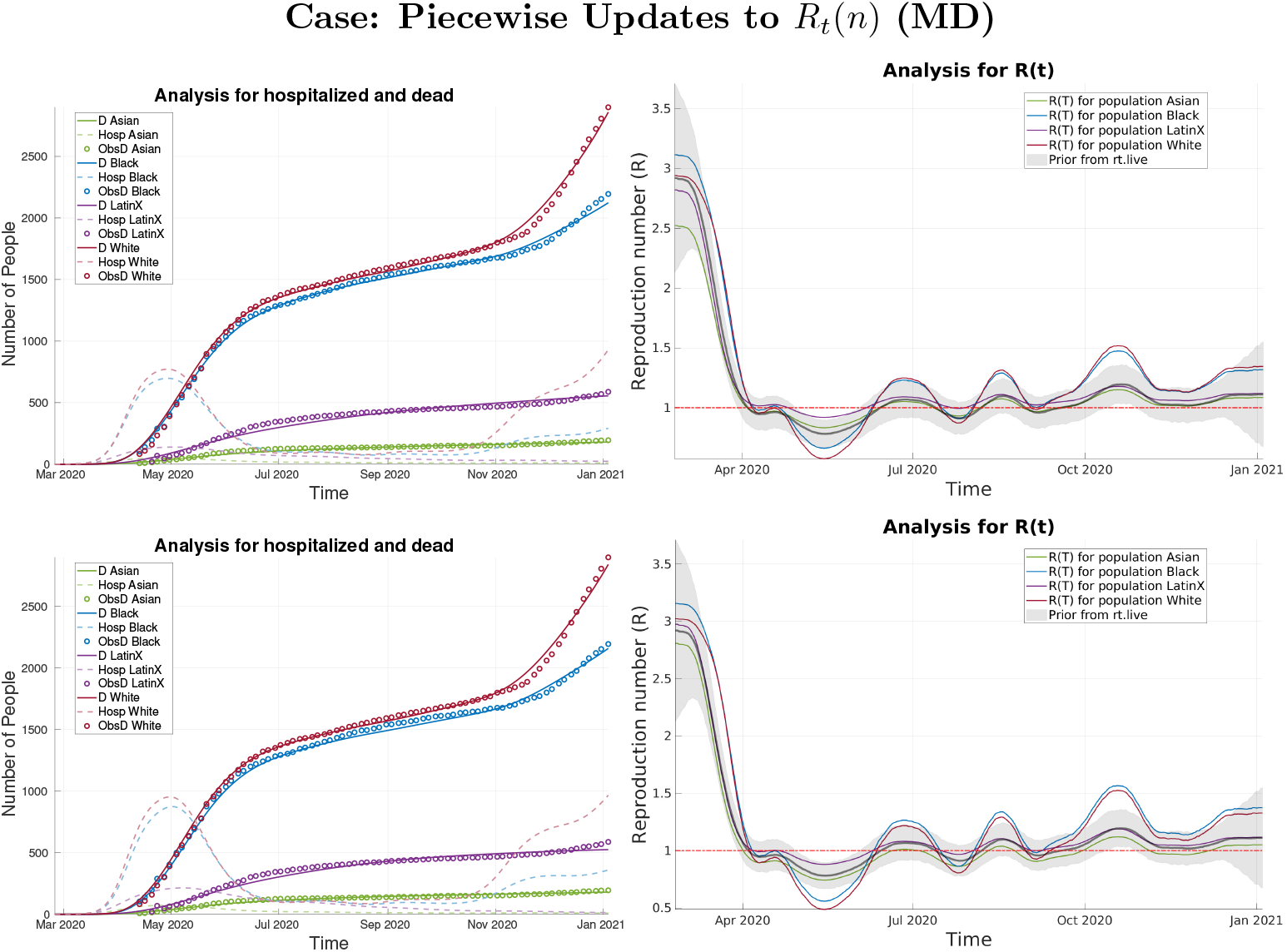
Analysis results when using peicewise updates to *R*(*t*) for MD. Top row: *R*^*A*^ with entries of all ones. Bottom Row: *R*^*A*^ for FLCs and NFLCs.

#### 4.2.8 The state of Michigan (MI)

The groups we consider for Michigan are shown in Table 3. In the state of Michigan, the Black community meets the criteria outlined in Section 4.2 to be an FLC. The Black population makes up 14% of the state’s population and comprises 24% of the deaths, while the White population makes up 78% of the state’s population and comprises 69% of the deaths. In all three run types, we see that during the intervention period the White population has the lowest values of the analysis *R*_*t*_(*n*) curves, again possibly signaling an ability to stay home due to the types of jobs held or as a result of reserve wealth. This is shown in Figures 25(Appendix B), and 15. We also see that early on in the pandemic, the Black and White populations have comparable numbers of infections despite the difference in population proportions. This is also shown in the aforementioned figures in the analysis of the exposed and infected as well as the recovered and total cases plots. We can also observe that for all three run types, in the same figures, that the Black community maintains a higher value of *R*_*t*_(*n*) for the vast majority of the data window.

**Figure 25.**
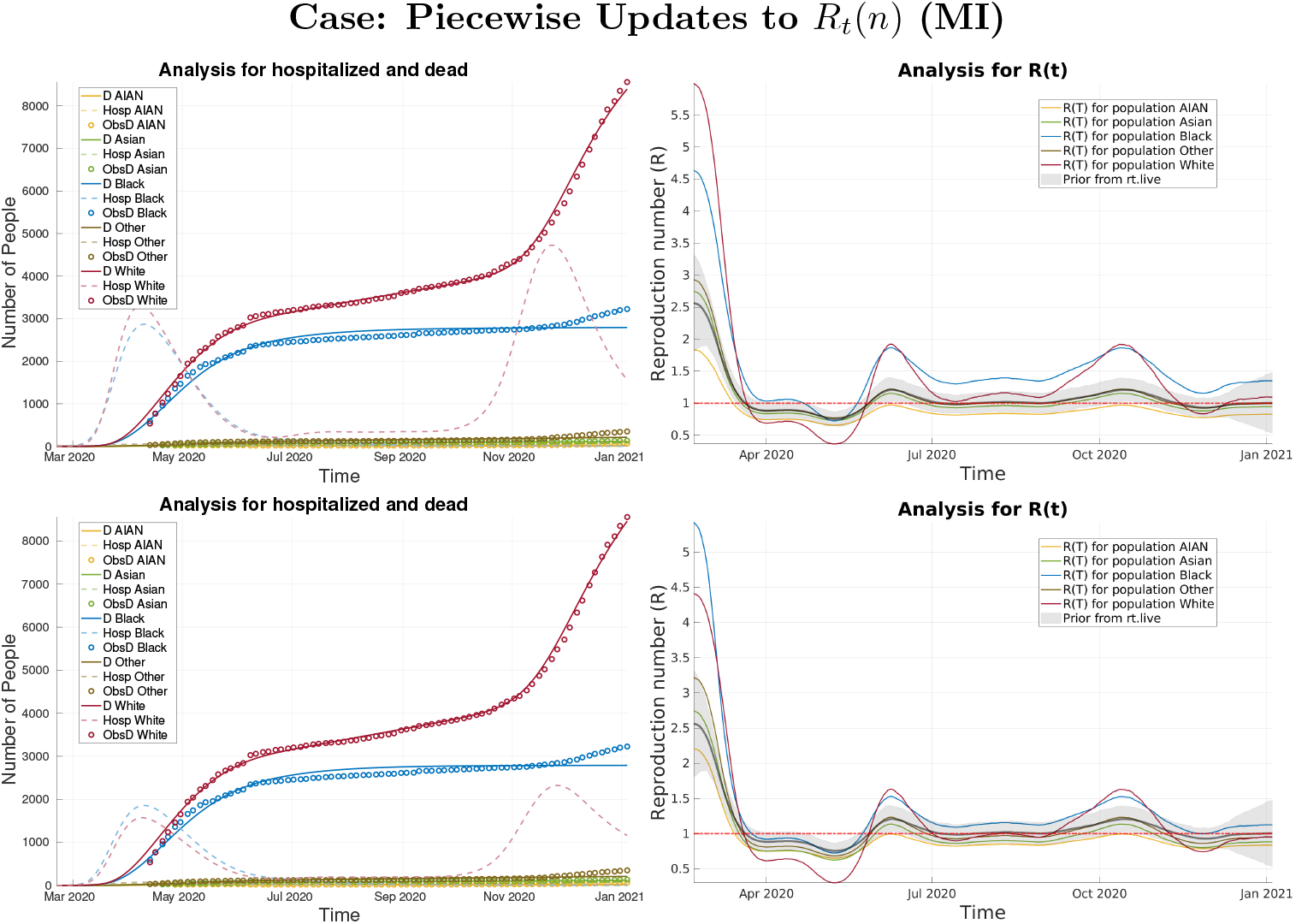
Analysis results when using peicewise updates to *R*(*t*) for MI. Top row: *R*^*A*^ with entries of all ones. Bottom Row: *R*^*A*^ for FLCs and NFLCs.

#### 4.2.9 The state of Utah (UT)

The groups we consider for Utah are shown in Table 3. In the state of Utah, none of the groups reported on meet the criteria to be an FLC as described in 4.2. In Figure 26 (Appendix B), and 16, we see a stratification of the analysis *R*_*t*_(*n*) curves amongst the groups with the White and Asian populations usually having the lowest values compared to other groups. In the analysis plot for *R*_*t*_(*n*) in the time-continuous case, we see a striking rise in the reproductive rate beginning at the end of the intervention period for the AIAN population (see Figure 16). Utah has a sizable amount of Native American reservation land, a group that was shown to be disproportionately affected by the pandemic. This spike is less apparent in the cases with piece-wise updates to *R*_*t*_(*n*), and we note though that the time-continuous update runs are better able to track time-dependent differences in the spread between groups. We also see a large spike in the continuous case at the end of the summer for the Other group. Also interesting here is that the disparity between different groups during the intervention period is somewhat less apparent than in other states. One possible reason for this is the large population of members of the Latter Day Saints (LDS) faith. The LDS church often offers financial and food assistance to its members. This could act as a cushion for LDS members of the Latinx community who were ineligible to receive assistance from the C.A.R.E.S. act allowing them to remain home longer.

**Figure 26.**
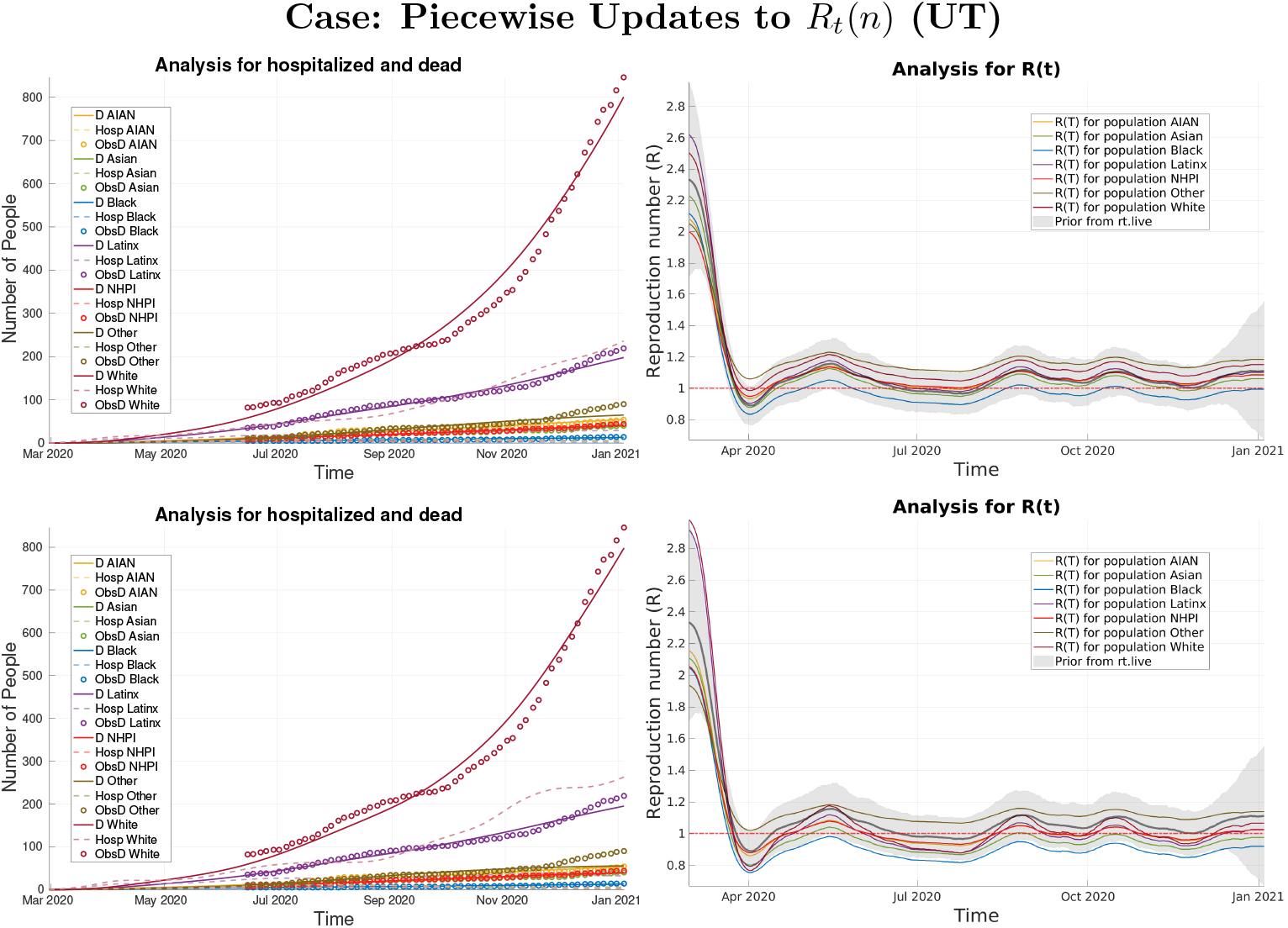
Analysis results when using peicewise updates to *R*(*t*) for UT. Top row: *R*^*A*^ with entries of all ones. Bottom Row: *R*^*A*^ for FLCs and NFLCs.

#### 4.2.10 The state of Washington (WA)

The groups we consider for the state of Washington are shown in Table 3. In Washington, the Latinx, NHPI, and AIAN populations meet the criteria for an FLC, as described in Section 4.2. The Latinx community comprises 13% of the total population with 34% of the confirmed cases, the NHPI community is less than 1% of the total population yet makes up 2% of the deaths, and the AIAN community comprises 1% of the population while accounting for 2% of the deaths. The disparity for the AIAN and NHPI communities is evidenced in all three run types with their analysis *R*_*t*_(*n*) values predominately above *R* = 1 in all three cases shown (Figures 27 (Appendix B) and 17). We also observe in the analysis for *R*_*t*_(*n*) plots in the continuous update case (Figure 17) that the Latinx community has higher values for the reproductive rate than all other groups–except for the NHPI and AIAN populations–during the intervention period.

**Figure 27.**
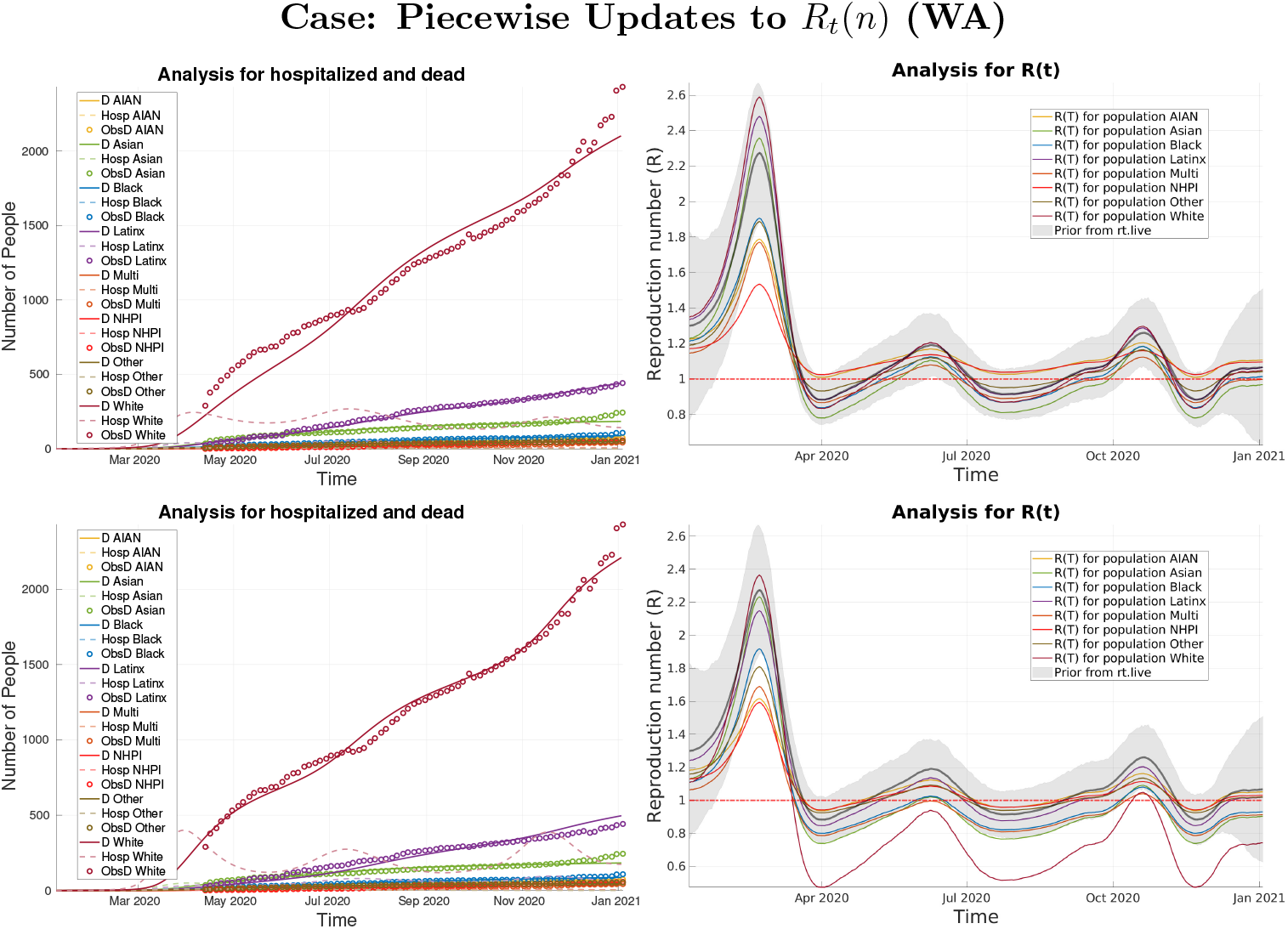
Analysis results when using peicewise updates to *R*(*t*) for WA. Top row: *R*^*A*^ with entries of all ones. Bottom Row: *R*^*A*^ for FLCs and NFLCs.

## 5. Conclusion and Discussion

We have developed a multi-population SEIR model with age classes and employed an ESMDA scheme to perform an initial re- analysis of the spread of SARS-CoV2 among different racial/ethnic groups. We find disparities in the rate of spread for different groups and estimate that rate as a function of time. We believe the primary driver of these disparities is the abilities of these groups to self-quarantine and avoid exposure. Factors that impact one’s ability to self-quarantine include an individual’s type of employment (work from home or not), access to reserve funds such as generational wealth, and access to healthcare or the general infrastructure in their specific locality. Groups for which many members cannot work from home or do not have reserve funds naturally will be exposed more often to the disease, eventually bringing the virus home to their family and close friends group where spread happens robustly and rapidly [22].

We think of these groups as frontline communities (FLCs). In our analyses, we find that typically the Black, Latinx, American Indian and Alaskan Native (AIAN), and Native Hawaiian and Pacific Islander (NHPI) populations exhibit *R*_*t*_(*n*) curves suggestive that they fit the criteria to be considered an FLC. This is typically most notable during and following the intervention period (lockdown). For the Latinx community, many were ineligible to receive financial assistance from the C.A.R.E.S. act due to immigration status making isolation at home during these periods difficult. Stimulus funds may also be less deliverable to FLCs, such as the Black community, which is often underbanked complicating fund distribution. After these intervention periods, FLCs risked increased exposure by returning to work in jobs that cannot be done from home. We consistently find that the White population, while a larger proportion of the overall population, had lower *R*_*t*_(*n*) values until the beginning of the Fall surge around October. This suggests that this group was better able to self isolate for a far longer period of time than the other groups we consider. This is consistent with what is known about wealth disparities between the White population and minority groups in the United States [27].

In some cases, we find evidence that a group is an FLC even though they do not fit the specific criteria outlined in Section 1.1. This is the case in the state of Connecticut where we see evidence that the Latinx community meets the criteria to be an FLC in terms of a disproportionate number of confirmed cases while the Black community does not. However, the analysis suggests more infections among the Black community than the Latinx community, despite being a smaller percentage of the population. This is based on the assimilation of the number of deaths and suggests that the Black population in the state may have poor access to testing or are less likely to seek a test. With the time-continuous analysis, we can also detect when spread began for a subgroup. We note that in the state of Alaska, spread begins in the White population and later into the AIAN population. This suggests major cities like Anchorage were hit first then spread to the rural regions later. The disparity in the number of deaths between the majority White population and the AIAN population also suggests how important healthcare infrastructure is in reducing the impact of the virus. Many of the far north rural communities in Alaska have little healthcare infrastructure and deliveries of supplies can be extremely difficult. We argue that lessons can be learned from these types of analyses to improve planning for a future pandemic.

Our main goals in this work were to study disparity in the spread of SARS- Cov2 and to demonstrate the utility of models and data assimilation to aid in understanding how the pandemic evolved among different racial/ethnic groups in various regions of the United States. This kind of analysis can provide important insight into successes and failures of policy as well as highlight causes for disparities. Using techniques such as ESMDA can allow us to “fill in the data gaps” presented when only considering things like confirmed cases or looking at general statistics. Armed with more complete information, better policy, and more effective planning can be implemented to help avoid such disparities and reduce the loss of life in a future pandemic. We note that at this stage the data available is a bit sparse and somewhat incomplete. While we make some conjectures as to the causes of disparities we find in our analyses, we stress that direct causation is not assured. However, as the data is better curated in the coming months and years, analyses such as these can be a powerful tool for social scientists, epidemiologists, and other experts to understand how and why events unfolded as they did and to find better ways to prepare in the future.

## Data Availability

The data was obtained from the covid tracking project.
https://covidtracking.com/

## 6. Acknowledgments

The authors would like to thank the AIM & MCRN summer program (2020) for their support in supervising, guiding, and helping fund this project. In particular, our mentors Nancy Rodriguez at CU Boulder and Chris Jones at UNC-Chapel Hill for their guidance during the summer program. We would like to also acknowlege the frontline communities and frontline workers who keep society functioning at the risk of their own lives. C. Sampson was supported by the US Office of Naval Research under grant N00014-18-1-2204. Daniel P. Maes was supported by the NSF GRFP DGE 1256260 grant. G. Evensen was supported by internal funding from NORCE.

## Appendix A. Model Parameter Priors

In Table 4 we show the first guess parameters used for all communities and in Table 5 we show the p-factors and CFR initially chosen and used for all communities.

## Appendix B. Piece-Wise Assimilation

Here we present the analysis for hospitalized and dead as well as *R*_*t*_(*n*) when applying piecewise updates to *R*_*t*_(*n*) in the cases that 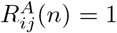 and when using our age stratified matrices for the FLCs and NFLCs discussed in Section 4.1.

